# Bench-to-bedside translation of Self-Healing colloidal hydrogels as 2^nd^ generation design of Flowable Hemostatic Matrix: From Preclinical evaluation to Human Clinical Trials

**DOI:** 10.1101/2025.11.27.25338626

**Authors:** Ganjun Feng, Kaiwen Chen, Qiwei Ying, Shuya Wang, Xin Li, Guanlin Li, Yantao Zhao, Binghua Ma, Huanan Wang, Dawei Li, Lijun Zhang, Jian Zhang, Yu Zhao, Changle Ren, Baodong Chen, Yueming Song

## Abstract

The emergence of self-healing materials has brought a paradigm shift in the design of functional biomedical devices for applications such as drug delivery, tissue regeneration, and 3D bioprinting. However, their clinical translation remains limited due to challenges including insufficient mechanical strength, potentially cytotoxic chemical modifications, and complex healing activation conditions. Herein, we present the development of self-healing colloidal gelatin hydrogel as an innovative design of flowable hemostatic matrix, and successfully demonstrate its bench-to-bedside translation into a biomedical device (named as Colloidose^®^). Specifically, amphoteric gelatin submicron particles self-assemble into an integrated gel network exhibiting a high storage modulus (G’> 15 kPa) and a healing efficiency exceeding 95%, enabling rapid in situ solidification to accelerate blood clot formation. By comparing with more conventional design strategy flowable gelatin matrix based on dispersion of hundreds micrometer-sized gelatin granules, we demonstrate that Colloidose^®^ are more effective for hemostasis in anatomically challenging or pressure-intolerant sites such as hepatobiliary surgery, otorhinolaryngology, and gynecology based on comprehensive preclinical animal studies and over 300 clinical cases. Overall, Colloidose^®^ exemplifies the successful clinical translation of an advanced self-healing biomaterial, establishing its role as a 2^nd^ generation flowable hemostatic matrix and opening new avenues for the development of injectable and moldable biomedical devices.

## 1 Introduction

Given the potentially devastating consequences, achieving adequate hemostasis is critical in all surgical procedures. Diffuse oozing hemorrhage from cavity walls of organs such as the liver or kidneys can result in substantial blood loss, prolonged operative times, and increased risks of infection and mortality^1, 2^. Moreover, hemorrhage can obscure visualization within confined operative fields, particularly during endoscopic procedures^3, 4^. Consequently, effective hemostatic strategies are essential for managing blood loss, mitigating hemorrhagic complications, and reducing transfusion requirements. Conventional clinical hemostasis methods—including vascular electrocauterization and suturing—are constrained by prolonged procedure duration, limited efficacy in emergency scenarios, suboptimal hemostatic outcomes, and potential collateral tissue damage^5–7^. Preformed hemostatic biomaterials (e.g., gauze, sponges) face limitations such as compromised deliverability through minimally invasive approaches and inadequate interfacial conformability with wound surfaces, restricting their utility for complex intracorporeal hemorrhage^8, 9^. By comparison, topical hemostatic agents (tissue adhesives and flowable matrix) offer superior applicability in intricate scenarios due to their injectability and tissue adaptability^10, 11^. These function either through chemical bonding with tissues or local delivery of pro-coagulants. However, hemostasis via chemical reactions or coagulation cannot achieve immediate hemorrhage control due to inherently gradual polymerization or clot formation^12, 13^. For example, pre-polymerized precursors used in adhesives (e.g., PEG-based sealants, fibrin glues) remain fluidic at the time of application, impeding stable localization at active bleeding sites. Once polymerized, these materials become rigid and non-reversible, and any mismatch between the cured matrix and the tissue surface may result in ineffective sealing. Current flowable hemostatic agents, including Surgiflo™ and Floseal™, consist of aggregated microparticle-based matrices and are clinically indicated for bleeding control in anatomically complex or compression-intolerant sites. Nonetheless, their relatively low mechanical strength and susceptibility to disintegration can limit efficacy, especially under high-flow or high-pressure bleeding conditions where material displacement may occur^14^. Therefore, the development of flowable hemostatic materials that integrate minimally invasive delivery, tissue conformity, and robust mechanical integrity remains a critical goal in surgical hemostasis.

Self-healing hydrogels based on reversible bonds have emerged as materials combining flowability and mechanical stability due to their thixotropic nature, which enables shear-induced liquefaction and self-solidification under static conditions^15, 16^. However, conventional polymeric self-healing hydrogels suffer from slow healing rates (minutes to hours), strict healing requirements (specific temperature/pH), and poor biocompatibility due to potentially cytotoxic chemical modifications, limiting their biomedical applications^17^. To address these challenges, our previous studies have demonstraed the conception of colloid-asembled self-healing (CASH) hydrogels that are composed of reversibly crosslinked particulate networks dispersed in aqueous phases^18, 19^. These CASH systems show promise for drug delivery, bioelectronics, and injectable wound dressings owing to their unique microarchitecture and viscoelastic behavior^20^. Critically, compared to conventional polymer self-healing hydrogels, CASH gels exhibit faster healing rates and superior mechanical strength due to versatile material selection and rapid particle rearrangement during healing^21,22^. Moreover, CASH gels are typically shear-thinning to enable injection/extrusion but also capable to solidify immediately upon release the stress, thereby allowing applications as injectable and moldable matirx that can be deliered via minimal or non-invasive way. Based on these superior properties of CASH gels, we suggested they can be utilized as effective flowable hemostatic matrices, which can be delivered via injection at the internal bleeding site, and thereby triggering sol-gel transition to enable local adaptation to the irregularly shaped defects, and form a physical barrier to block the wound and prompt hemorrhage control. Validation of this approach could enable the development of a high-strength and flowable hemostatic matrix with enhanced clinical utility.

Based on this CASH strategy, we herein developed gelatin-based colloidal gels (commercially named Colloidose^®^, supplied by HUANOVA Biotech.) as a second-generation flowable hemostatic matrix for rapid and robust sealing across diverse tissue types. This system consists of amphoteric gelatin submicron particles that undergo spontaneous bottom-up assembly via nanoparticle coalescence, forming a physically crosslinked, interconnected colloidal network. The non-covalent interactions among nanoparticles endow the gel with shear-thinning behavior and self-healing capacity, enabling minimally invasive delivery while preserving high mechanical integrity. To assess its performance, the hemostatic potential of CASH gels was evaluated through rheological characterization, in vitro coagulation testing (including thromboelastography, TEG), and in vivo degradation and stability assessments. A commercially available flowable hemostatic gelatin matrix (Surgiflo^TM^, Johnson & Johnson, USA), composed of micron-scale porous gelatin particles, was used as the clinical benchmark for comparison. Results demonstrated that CASH gels exhibited superior mechanical strength, enhanced tissue adhesion, and improved retention in fluid environments compared to the microstructured reference, all of which are critical attributes for effective hemostatic sealing. Moreover, the clinical relevance of Colloidose^®^ was confirmed through large-animal studies and multicenter human trials, where it demonstrated efficacy in managing bleeding in pressure-sensitive or surgically restricted regions, including neurosurgery, spine, cardiothoracic surgery, hepatobiliary interventions, otorhinolaryngology, and gynecology. Overall, our CASH gel platform exemplifies the clinical translation of a next-generation self-healing biomaterial, consolidating its role as a second-generation flowable hemostatic matrix with broad translational potential for modern surgical applications.

**Scheme 1.**
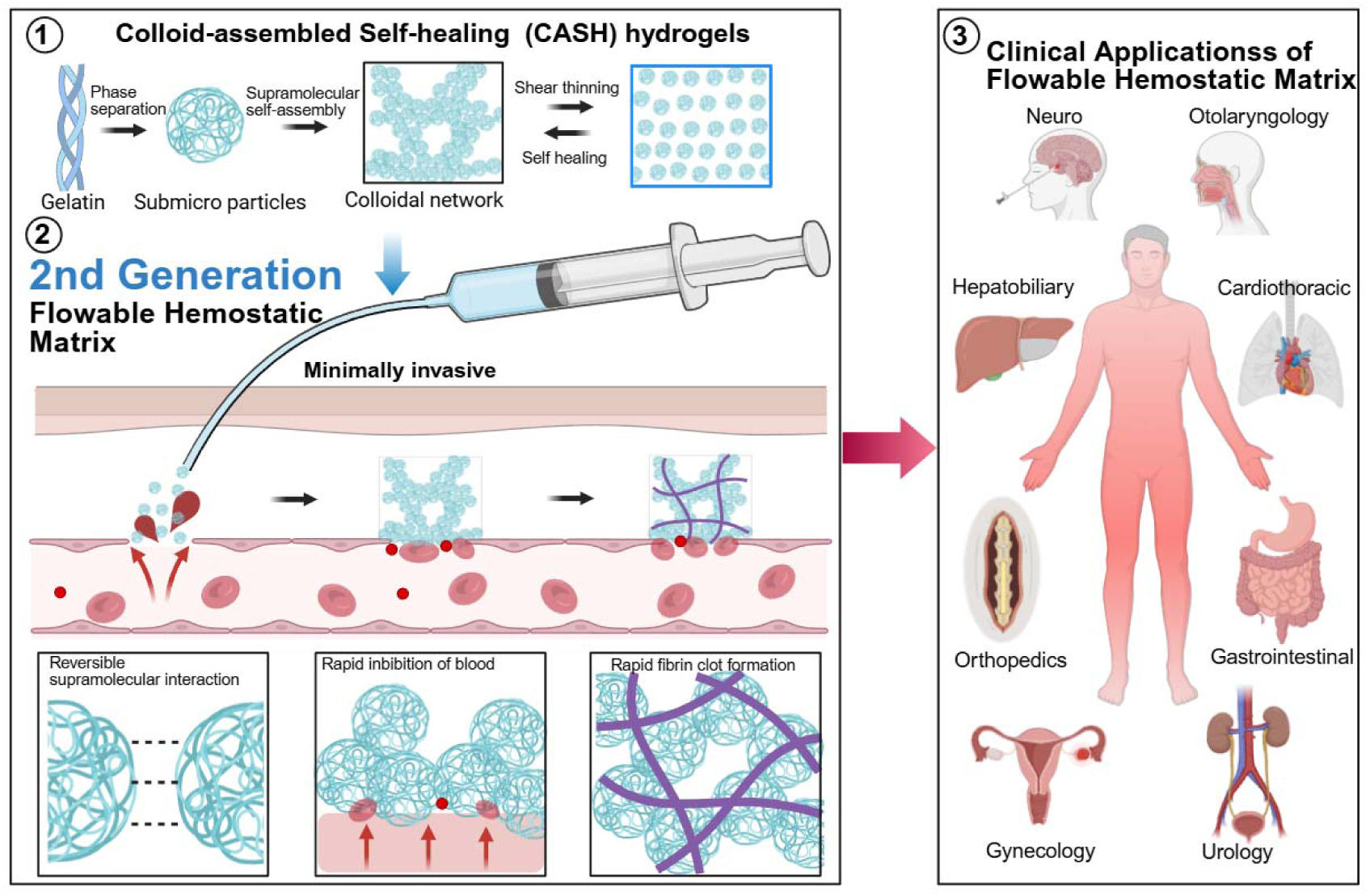
Schematic illustration showing the broad clinical applications of flowable hemostatic matrices across multiple surgical fields and the design of CASH gels as a second-generation flowable hemostatic matrix. CASH hydrogels are constructed via supramolecular self-assembly of gelatin submicroparticles into a dynamic colloidal network, conferring shear-thinning injectability and autonomous self-healing. Upon minimally invasive delivery, the hydrogel rapidly fills bleeding sites, inhibits blood loss, and accelerates fibrin clot formation, thereby achieving rapid hemostasis and preventing rebleeding in diverse clinical scenarios.

## 2 Result and discussion

### 2.1 CASH gels possess distinct structural and compositional features as a flowable hemostatic matrix

To compare the chemical compositions of the gelatin-based CASH hydrogel and the commercially available microgranular gelatin-based hemostatic matrix (Surgiflo™), Fourier-transform infrared (FT-IR) spectroscopy was conducted. The FT-IR spectra of both materials exhibited characteristic proteinaceous absorption bands, including the amide II band (N-H bending coupled with C–N stretching, 1530 to 1590 cm^-1^) and the amide III band (N-H and C-N planar deformation, 1200 to 1250 cm^-1^), which are consistent with their gelatin-based composition. Notably, Surgiflo™ showed a slight blue shift and attenuation of the N-H stretching band, which suggests the presence of enhanced intermolecular hydrogen bonding resulting from its thermal cross-linking process. In contrast, CASH presented a distinct peak at 1340 cm^-1^, which was not observed in Surgiflo™ and is attributed to glutaraldehyde-derived Schiff-base linkages that form cyclic pyridine structures. Quantitative analysis of secondary structures based on infrared peak deconvolution and integration revealed that CASH contains 45.3% β-sheet, 43.2% α-helix, and 11.5% β-turn conformations. In comparison, Surgiflo™ contains 35.8% β-sheet, 45.0% α-helix, and 19.1% β-turn, as shown in Figure 1b. The higher β-sheet content in CASH likely results from its bovine-derived protein source, which is inherently richer in β-sheets than porcine gelatin, and from structural reorganization induced by nanoparticle assembly. These features may contribute to greater molecular stability^23, 24^. We further characterized the microstructures of CASH and Surgiflo™ using scanning electron microscopy (SEM). SEM imaging revealed that CASH consisted of highly refined, uniform nanoscale particles (average diameter 293.4 ± 133.4 nm) that serve as building blocks and assemble into an interconnected particulate network^25–27^. In contrast, Surgiflo™ was composed of micrometer-sized porous gelatin granules with an average diameter of ≈600 μm (Figure 1e)^28–30^. When these granules pack to form an integrity, they accumulate into a loose, porous, packed particulate network in which individual particle boundaries become indistinct. By comparison, the CASH gel forms a densely packed nanocolloidal network with a refined hierarchical architecture (Figure S1). This contrast highlights a fundamental difference in material architecture. Traditional flowable matrices rely on loosely packed microparticles held together by weak intergranular forces. In contrast, CASH forms a continuous nanoparticle network stabilized by stronger interparticle attractions, resulting in greater structural integrity and coherence (Figure 2d,f).

**Figure 1.**
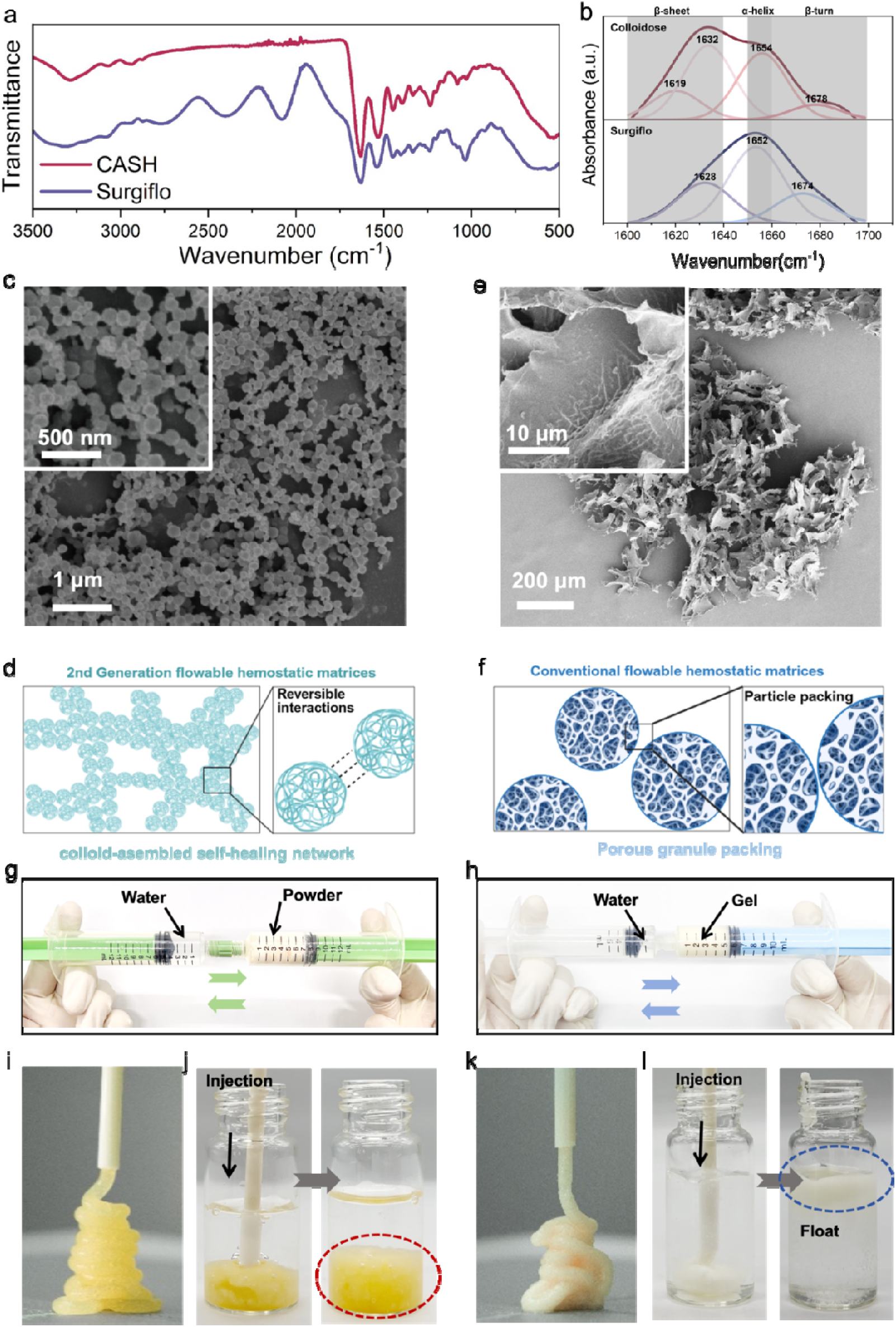
(a) FT-IR spectra of freeze-dried CASH gels and Surgiflo™, both showing characteristic proteinaceous absorption bands of gelatin. (b) Second-derivative profiles reveal additional peaks in CASH gels associated with glutaraldehyde-mediated Schiff base crosslinking, whereas Surgiflo™ displays signatures of thermal denaturation. (c) Scanning electron micrograph of CASH gels demonstrating a densely interconnected nanoscale particulate network (average particle size ≈ 300 nm) forming a compact hierarchical framework. (d) Schematic of CASH gel preparation, involving electrostatic assembly of gelatin nanospheres stabilized by glutaraldehyde crosslinking. (e) By contrast, Surgiflo™ exhibits loosely packed microspheres on the micrometer scale (≈ 600 μm), resulting in a porous and mechanically fragile structure. (f) Schematic of Surgiflo™ preparation through thermal processing of microgranular gelatin. (g) Optical image of CASH gels illustrating smooth injectability through a standard syringe and high moldability for defect-conforming application. (h) Optical image of Colloidose^®^ showing excellent aqueous stability under physiological conditions. Optical images of CASH(i,j) and Surgiflo™(k,l) under identical conditions, demonstrating limited moldability and rapid disintegration with loss of structural stability.

**Figure 2.**
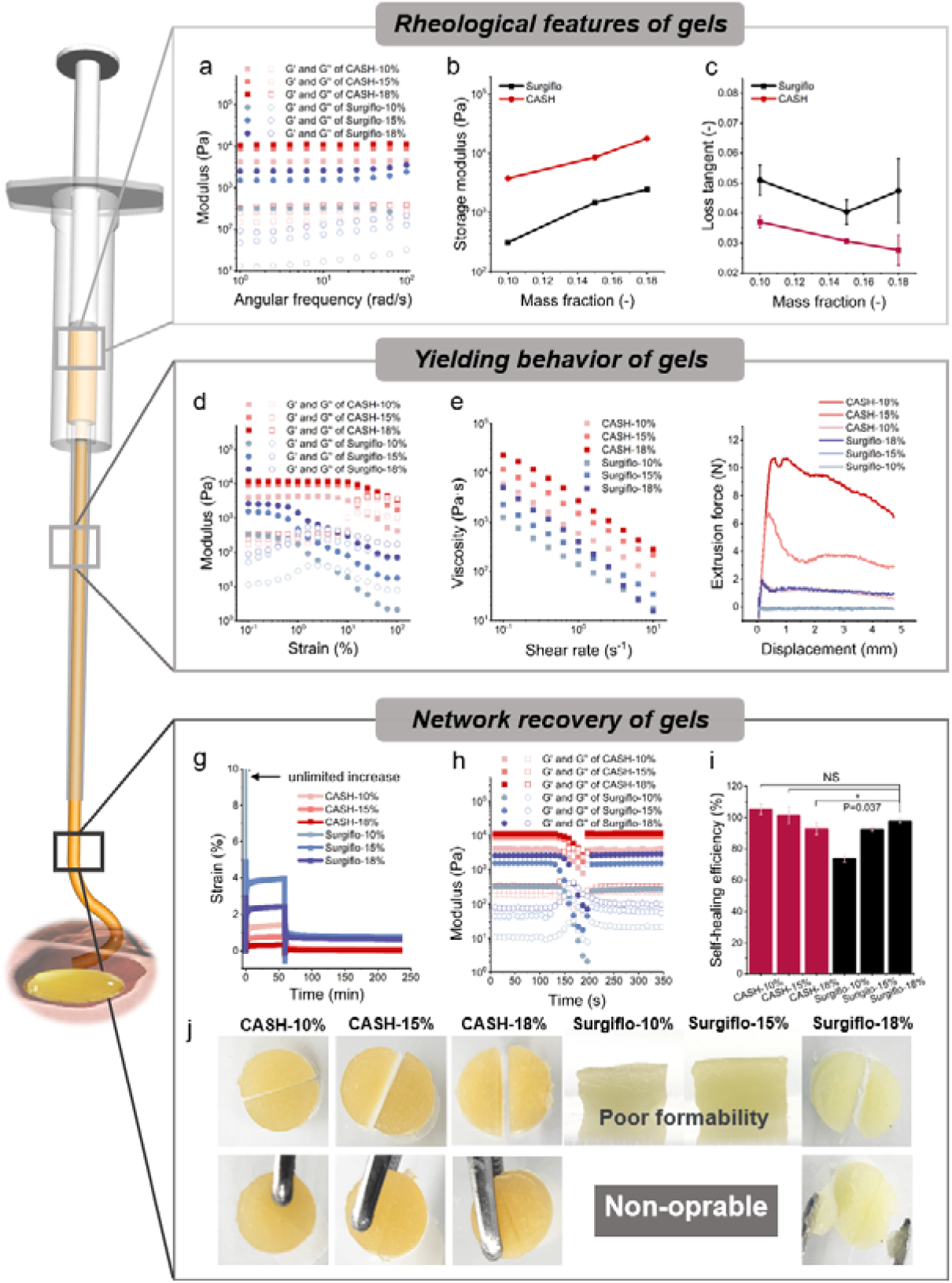
Rheological performance, injectability, and self-healing behavior of nanostructured CASH gels compared with microstructured granular gels (Surgiflo™) at different concentrations (10, 15, and 18 w/v%). (a) Frequency-dependent oscillatory measurements (0.1–100 Hz, strain 0.5%) showing that CASH gels consistently exhibit higher storage modulus (G’) and more stable network elasticity than Surgiflo™. (b) Dependence of G’ on concentration, confirming superior mechanical reinforcement of CASH gels across all tested mass fractions. (c) Loss tangent (tan δ, 1 Hz) indicating a more solid-like response for CASH gels compared with the viscoelastic-dominated Surgiflo™. (d) Strain-dependent G’ and G” (6.28 rad/s), demonstrating greater yield resistance and structural integrity in CASH gels. (e) Shear-thinning behavior showing rapid viscosity reduction under increasing shear rates, reflecting smooth injectability of both gel systems. (f) Extrusion force measurements during syringe injection, demonstrating lower injection resistance for CASH gels compared with granular matrices, thereby confirming their clinical operability. (g) Creep–recovery profiles under constant stress (10 Pa) and subsequent release (0 Pa), showing minimal residual strain in CASH gels, while granular gels undergo irreversible deformation. (h) Recovery of G’ following destructive shearing (0.1–1000% strain, 1 Hz), highlighting rapid and near-complete restoration of mechanical integrity in CASH gels. (i) Quantitative self-healing efficiency is defined as the recovered G’ relative to the initial value, confirming that CASH gels achieve significantly higher recovery compared with Surgiflo™ after injection-induced shear. (j) Photographs showing the self-healing behavior of the CASH gel.

CASH and Surgiflo™ were prepared according to the respective manufacturers’ protocols to evaluate key physical characteristics, including injectability, moldability, and mechanical stability. Surgiflo™ contains approximately 18% (w/v) gelatin microparticles in a gel-like base, while Colloidose^®^ comprises ∼10% (w/v) CASH gel powder. Both products are provided in sterile dual-syringe systems with Luer-lock connectors to allow intraoperative mixing and rapid gel formation. The Colloidose^®^ formulation is supplied as a dry yellow powder, whereas Surgiflo™ is pre-hydrated, appearing as a pale-yellow gel. Due to its water-containing formulation, Surgiflo™ is susceptible to evaporation and thermal denaturation, requiring controlled storage conditions (4∼25 °C). In contrast, the powder-based formulation of Colloidose^®^ is more thermally stable, eliminating the need for cold-chain storage and offering an extended shelf life.

To assess minimally invasive delivery performance, both materials were extruded through laparoscopic catheters. Optical imaging confirmed that both could be successfully delivered through narrow lumens. Upon extrusion, Colloidose^®^ formed cohesive, continuous strands with preserved shape fidelity (Figure 2i), whereas Surgiflo™ exhibited heterogeneous extrusion behavior, showing visible phase separation between gelatin and aqueous components, which led to discontinuous clumping. To simulate bleeding conditions, both gels were injected into phosphate-buffered saline (PBS). Colloidose^®^ adhered conformally to the container bottom and maintained structural cohesion (Figure 2j). In contrast, Surgiflo™ dispersed into floating fragments without preserving gel integrity. Furthermore, swelling analysis under aqueous conditions revealed that Colloidose^®^ expanded by less than 10%, indicating its dimensional stability in fluid environments. Due to the gel’s poor cohesion, this measurement was not feasible for Surgiflo™. These observations demonstrate that the nanoparticle-based colloidal network in Colloidose^®^ imparts a unique combination of injectability, conformability, aqueous stability, and low-swelling ratio, which are highly desirable for maintaining effective hemostatic function in dynamic, fluid-exposed surgical environments.

### 2.2 CASH gels exhibit stable rheological properties with injectability, self-healability, and high mechanical strength

Viscoelasticity is a critical parameter for flowable hemostatic matrices, as it governs their ability to conform to wound geometry and exert compressive sealing forces. We systematically characterized the viscoelastic behavior of CASH gels (Colloidose^®^) and Surgiflo™ at clinically relevant solid contents of 10%, 15%, and 18% (w/v), denoted as CASH-10%, CASH-15%, and CASH-18%, respectively, using oscillatory rheological measurements (Figure 2). To this end, oscillatory frequency sweeps were performed, which revealed minimal frequency dependence for CASH gels at all concentrations, as evidenced by the storage modulus G’ showing negligible increases with rising frequency (Figure 2a). In addition, the G’ values of CASH increased significantly from 4.9 ± 0.4 kPa to 18.0 ± 0.6 kPa as the solid concentration rose from 10 to 18% (w/v). By contrast, Surgiflo™ composed of micrometer-sized gelatin granules, exhibited a stronger frequency dependence in their viscoelastic response, with G’ showing a more pronounced dependence on frequency. Moreover, as the solid concentration increased from 10 to 18% (w/v), the granular gels presented G’values ranging from 0.31±0.05 kPa to 2.58±0.19 kPa—nearly an order of magnitude lower than those of the nanostructured CASH gels (Figure 2b). Notably, 18% (w/v) Surgiflo™ achieved only G’ ≈ 2.58 kPa, below the value measured for 10% colloidal CASH gels. This mechanical disparity can be rationalized by scaling laws for colloidal-gel elasticity based on interparticle interactions, wherein gel network modulus can be predicted by GCφ^n^U /d^3^, where φ is the volume fraction, U is the interparticle attractive potential energy, and d is the particle diameter^31–33^. Compared to colloidal gels, granular gels exhibited larger particle diameters and reduced interparticle attractive potential, thereby leading to significantly higher gel elasticity for nanostructured CASH gels than that of granular gels.

Further analysis confirmed the superior elastic dominance of colloidal gels (CASH) over granular gels (Surgiflo™), as evidenced by lower loss factors (tan δ) at equivalent mass fractions (Figure 2c). This elastic behavior intensified progressively with increasing concentration from 10 to 18% (w/v), demonstrating a concentration-dependent mechanical reinforcement. Notably, CASH reached a close-packed state even at 10 %(w/v), in which amphoteric gelatin nanoparticles maximize interparticle contact points. These contact points enable collective non-covalent interactions, including electrostatic and hydrophobic forces, which govern the material mechanical response. In contrast, granular gels exhibited sparse particle contacts even at 18% (w/v) owing to micrometer-scale steric constraints. This contact deficiency limits synergistic bonding networks, thereby favoring viscous dissipation over elastic recovery. Although individual non-covalent bonds possess low binding energies, their concerted action establishes a stable gel network. Overall, CASH exhibits markedly higher mechanical modulus and elasticity, enabling the gel to exert effective compressive hemostatic action at the bleeding site.

The yield behavior of flowable hemostatic gels plays a critical role in ensuring their minimally invasive injectability. To assess this, we first conducted dynamic amplitude sweep tests (oscillatory strain sweeps) to evaluate shear deformation. Granular gels exhibited an approximately linear decline in G’, followed by yielding at strains of 6.8% ± 1.2% (10% w/v), 4.5% ± 0.8% (15% w/v), and 8.9% ± 1.5% (18% w/v) (Figure 2d). In contrast, CASH gels displayed a characteristic elastic-gel response, with minimal reduction in G’ (<5%) under low strain (<10%) and significantly higher yield strains ranging from 16.3% ± 2.1% (10% w/v) to 63.7% ± 5.3% (18% w/v). Furthermore, shear-thinning properties were quantified through steady-state shear tests. Both systems exhibited decreasing viscosity with increasing shear rate (Figure 2e), conforming to the power-law model τ=Kγ^n^, where τ is the shear stress, K is the consistency coefficient, γ is the shear rate, and *n* is the flow behavior index. Accordingly, the average *n* values were calculated to be −0.84 and −1.01 for the granular and colloidal gels, respectively. This indicates that the granular gels exhibit weaker shear-thinning than the colloidal gels, suggesting that the latter possess greater responsiveness to shear. Further injection-force tests showed that the more elastic colloidal gels required high plunger forces (< 10 N) for smooth extrusion, whereas the granular gels exhibited lower injection force consistent with their lower modulus (Figure 2f). Notably, CASH-10% required an injection force of 1.97 ± 0.15 N, comparable to Surgiflo^®^ (2.05 ± 0.18 N) despite its higher consistency index. Taken together, these results confirm pronounced shear-thinning and low injection forces for both systems, supporting their suitability for minimally invasive delivery.

Injection-induced shear thinning can transiently compromise the mechanical stability of flowable hemostatic gels, making rapid post-injection recovery of strength critical for achieving effective compressive hemostasis at wound sites. To assess this property, we first examined strain-recovery behavior using creep tests under a constant stress of 10 Pa. Granular gels exhibited progressive creep deformation, reaching strains of 3.97% ± 0.21% at 15% (w/v) and 2.42% ± 0.15% at 18% (w/v) after 630 s (n = 5). Recovery was incomplete, with residual strains of 0.76% ± 0.08% and 0.64% ± 0.07%, respectively (Figure 2g). Granular gels at 10%(w/v) surpassed their yield-stress threshold under the applied load, resulting in structural fluidization. This was evidenced by uncontrolled strain accumulation, reflecting irreversible network collapse caused by dissociation of dynamic bonds. In contrast, CASH gels displayed predominantly elastic behavior, with immediate strain responses of ∼1.46%, 0.77%, and 0.36% at 10%, 15%, and 18% (w/v), respectively, followed by stable strain plateaus over 60 s at 10 Pa. Upon removal of stress, CASH gels recovered almost completely, retaining no more than 0.04% residual deformation (Figure 2g).

We further evaluated the thixotropic behavior of the gels using a three-step oscillatory shear protocol, comparing CASH gels with the commercial Surgiflo™ system. This test involved assessing the mechanical moduli under initial undisturbed conditions (Region I), monitoring their evolution under continuous high-strain shear (Region II), and subsequently evaluating the recovery of viscoelastic properties once the shear stress was removed (Region III)^34, 35^. In Region I, both gels exhibited stable viscoelastic behavior under low strain (1%, 1 min), with storage modulus (G’) exceeding loss modulus (G”), indicating intact network structures. Upon applying destructive shear (Region II: 1–500% strain, 1 min), both systems underwent a gel-to-sol transition, reflecting disruption of their internal networks. Remarkably, the CASH gels exhibited rapid and efficient self-healing behavior. All tested groups recovered more than 90% of their initial storage modulus within seconds after cessation of shear (Region III), a level of performance attributed to the strong electrostatic interactions and hydrogen bonding among the densely packed gelatin nanoparticles, which promote quick reassembly of the colloidal network. By comparison, highly concentrated Surgiflo™ gels also demonstrated thixotropic recovery, with a measurable but lower extent of modulus restoration (typically less than 95%) following destructive shear. The high specific surface area of the porous gelatin microparticles in Surgiflo™ may facilitate interparticle cohesion during dense packing^36, 37^, contributing to its ability to partially restore mechanical integrity. However, the particulate network of Surgiflo™ exhibited limited self-healing efficiency under diluted conditions. This may be attributed to the reduced particle packing density upon dilution, which weakens interparticle interactions and hinders effective network reformation. This behavior was further confirmed by morphological observations (Figure S2), where exposure to excess aqueous environments led to collapse of the gel matrix, resulting in particle disassembly and dispersion. These findings suggest that while both materials exhibit thixotropic characteristics, the nanoparticle-based CASH system demonstrates superior network resilience and self-recovery capacity, particularly under dynamic physiological conditions where shear and dilution frequently occur.

Comparative evaluation of flowable hemostatic matrices revealed that both systems are readily injectable and can be reconstituted after extrusion. However, CASH gels extrude as continuous, defect-conforming strands that maintain structural integrity and minimal swelling (<10%), whereas the Porous Granule Packing (PGP) design, exemplified by Surgiflo™ and Floseal™, tends to fragment and phase-separate in aqueous environments (Figure 2). The PGP architecture represents the first-generation design paradigm of flowable hemostatic matrices, in which micrometer-scale gelatin granules are loosely packed within a carrier medium. This configuration facilitates injection but inherently limits interparticle contact density and cohesive strength, leading to poor mechanical stability and structural collapse under fluid exposure or high-pressure bleeding conditions ^38, 39^. In contrast, the CASH system introduces a second-generation colloidal design, where submicron gelatin nanoparticles undergo bottom-up assembly into a densely percolated network stabilized by reversible noncovalent interactions (electrostatic, hydrogen-bonding, and hydrophobic forces). This architecture yields markedly enhanced viscoelastic properties—manifested as a two-to tenfold increase in storage modulus (G′ ≈ 5–18 kPa), lower loss factors (tan δ < 0.3), higher yield strains, and rapid creep recovery, while retaining excellent shear-thinning behavior and low injection forces. Collectively, these findings highlight a paradigm shift from the PGP to the CASH design, transforming loosely packed granular assemblies into self-assembled, cohesive colloidal networks. By integrating injectability with structural resilience and self-healing capability, CASH establishes a second-generation framework for flowable hemostatic matrices that enables durable and conformal sealing even under dynamic, high-pressure bleeding conditions.

### 2.3 CASH gels exhibit strong hemostatic sealing and pro-coagulant activity

Based on a comprehensive assessment of clinical operability, particularly the requirements for injection force and wound-filling conformability, we further examined the in vitro hemostatic performance of CASH gels at two concentrations (CASH-10% and CASH-15%) using a custom-designed hemorrhage simulation model. Fresh porcine hearts were integrated into a pressure-monitored perfusion system and continuously circulated with freshly collected porcine blood (Figure 3a). Given the primary clinical use of flowable matrices in venous hemorrhage—where physiological pressures typically range from 1 to 3 kPa (8–22 mmHg)—an artificial blood circulation loop was established using a peristaltic pump with pressure-controlled flow modulation^40, 41^. A spindle-shaped defect measuring 10 mm × 5 mm was created in the ventricular wall to generate a standardized active bleeding wound. Blood flowed outward under controlled pressure, simulating a physiologically relevant hemorrhage scenario. The pressure-monitoring system enabled quantitative evaluation of each material’s ability to achieve hemostasis and effectively seal the wound. A target venous pressure of approximately 3 kPa was maintained to simulate a clinically relevant worst-case scenario and rigorously evaluate the wound-sealing capacity of the tested materials. Visual inspection revealed minimal blood extravasation from the injured heart following treatment with CASH-15%, whereas both CASH-10% and Surgiflo™ (with a gelatin mass fraction of approximately 18%) were less effective in preventing leakage under identical conditions (Figure 3b). Quantitative measurements of simulated vascular pressure further confirmed the superior sealing performance of CASH-15%, which sustained a pressure of 4.95 ± 0.83 kPa—exceeding the clinically relevant venous threshold of 3 kPa^42, 43^—compared to 2.65 ± 0.41 kPa for CASH-10% and 2.11 ± 0.25 kPa for Surgiflo™ (Figure 3c). We hypothesized that the enhanced sealing efficacy of CASH-15% can be attributed to the improved tissue adhesion and mechanical strength of the colloidal gels. To validate this hypothesis, the adhesive strength of each gel formulation was measured using lap-shear testing with porcine skin as the substrate. The results indicated that Colloidose^®^ exhibited approximately five times greater adhesion strength compared to Surgiflo™ (Figure 3d). This enhancement is likely due to the higher specific surface area and conformability of its nanoscale colloidal network, which enables intimate mechanical interlocking with the tissue microtopography.

**Figure 3.**
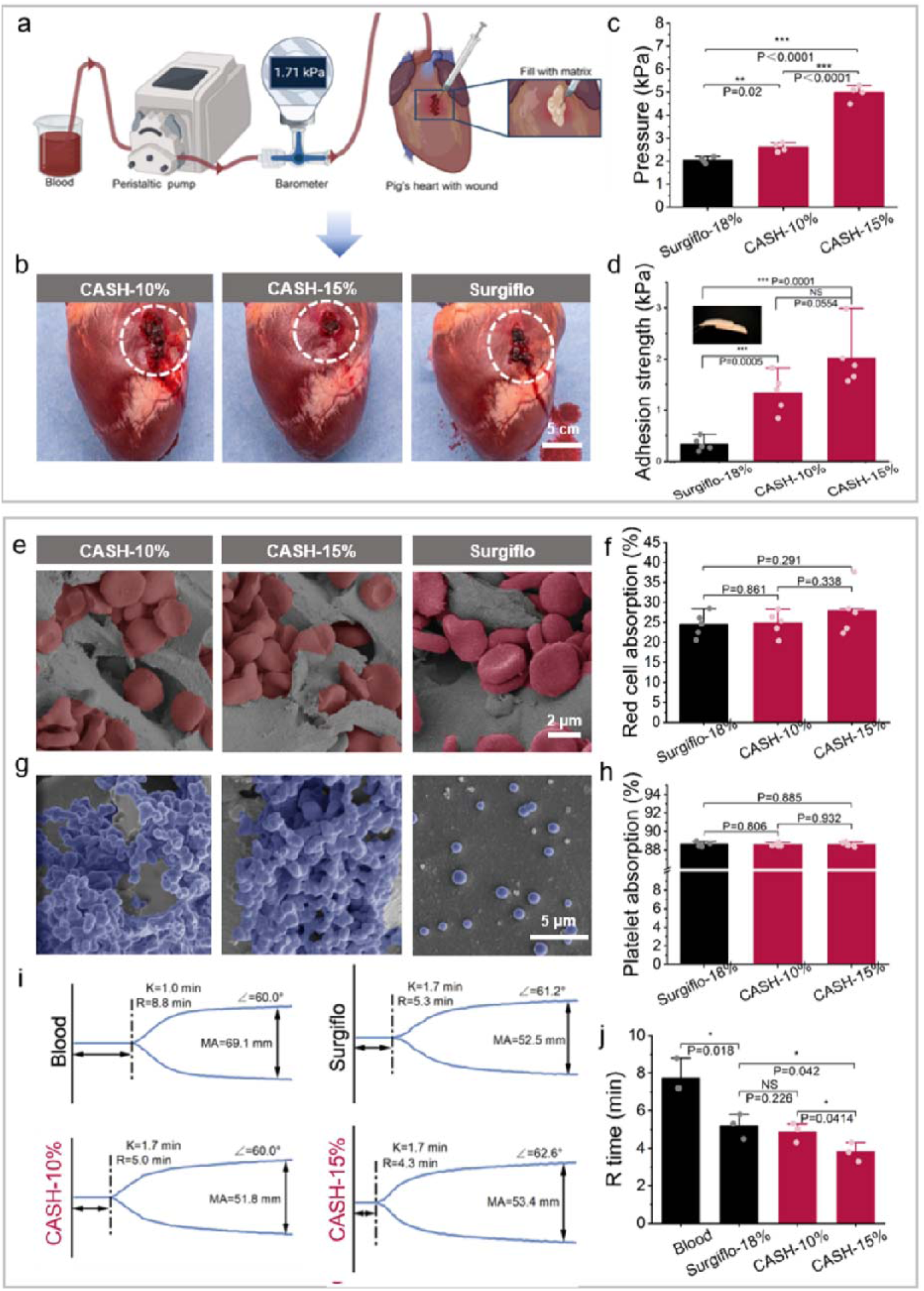
In vitro wound sealing efficacy and procoagulant activity of CASH gels compared with Surgiflo™. (a) Schematic of the custom perfusion setup for burst pressure testing, in which porcine hearts with standardized incisions were connected to a peristaltic pump to simulate venous pressure and monitored with a barometer. (b) Representative images of wounded porcine hearts treated with CASH-10%, CASH-15%, or Surgiflo™, demonstrating visible sealing of the defect area (dashed circle, scale bar: 5 cm). (c) Quantification of burst pressure, showing that CASH gels withstand significantly higher pressures than Surgiflo™, confirming superior sealing performance. (d) Adhesion strength measured by lap-shear testing on porcine tissue (1.5 × 1.0 cm overlap), demonstrating stronger interfacial bonding of CASH gels compared with Surgiflo™. (e) SEM images of red blood cells (RBCs) adhering (e) and platelet adhesion (f) to the surface of CASH gels and Surgiflo™. Quantitative analysis of RBC adsorption(g) and platelet adsorption(h) across CASH gels and Surgiflo™. (i) Representative thromboelastography (TEG) tracings of rabbit whole blood treated with CASH gels or Surgiflo™, highlighting accelerated clot formation kinetics for CASH gels. (j) R-time quantification derived from TEG analyses, showing significantly shortened clot initiation times for CASH gels compared with Surgiflo™ and untreated blod.

We next assessed the ability of the matrices to interact with blood cells by incubating them with red blood cell (RBC) suspensions and platelet-rich plasma (PRP). SEM imaging confirmed that both CASH gels and Surgiflo^TM^ could adsorb RBCs and platelets on their surfaces (Figure 3e, f). Quantitative analysis further demonstrated no significant differences in adsorption efficiency between the groups for either RBCs or platelets (Figure 3g). These findings indicate that CASH gels exhibit comparable blood cell adsorption capacity to Surgiflo^TM^, supporting their suitability for promoting coagulation and clot formation in hemostatic applications.

To further assess coagulation kinetics, thromboelastography (TEG) was performed using whole blood mixed 1:1 (v/v) with Surgiflo^TM^ or CASH formulations. Both materials significantly accelerated clot initiation compared with untreated blood, as reflected by reduced R-times—from 8.8±0.4 min (untreated) to 5.3±0.2 min (Surgiflo^TM^), 5.0±0.3 min (CASH-10%), and 4.3±0.2 min (CASH-15%) (Figure 3j). Notably, R-times achieved with CASH gels were significantly shorter than those observed with Surgiflo^TM^, confirming their superior procoagulant activity. This enhancement is likely attributable to the higher mechanical stability of CASH gels, which enables the formation of more robust blood-gel composites under shear and flow conditions (Figure 3i). Collectively, these results demonstrate that CASH gels support efficient cellular interactions and accelerate coagulation, thereby providing a mechanistic basis for their potent in vivo hemostatic performance.

### 2.4 CASH gels can serve as scaffolds for cell growth but also as carriers for sustained drug release

One key advantage of gelatin-based hemostatic matrix lies in their inherent cytocompatibility, which supports cell adhesion and may promote subsequent tissue ingrowth and regeneration at wound sites. To assess this potential, CASH and Surgiflo™ scaffolds were molded and immersed in culture medium. CASH maintained structural integrity for several days, whereas Surgiflo™ disassembled within hours, suggesting that particulate architectures composed of large micrometer-sized granules are more prone to dispersion in vivo and therefore less capable of providing sustained structural support during healing. Such instability also limits their applicability as cell-laden scaffolds for regenerative purposes.

Since Surgiflo™ rapidly disassembled and could not maintain scaffold integrity under culture conditions, only the cytocompatibility of CASH gels was further assessed in a two-dimensional cell culture model. MC3T3 cells were seeded on CASH-10% and -15% gels and evaluated using CCK-8 and live/dead assays. Both formulations significantly enhanced cell proliferation after 24 h, with comparable viability maintained at Days 1, 3, and 5. Live/dead staining revealed predominantly spindle-shaped viable cells with extensive pseudopodia, while immunofluorescence of α-actinin demonstrated well-organized cytoskeletal structures (Figure 4 c–e). These findings confirm not only the excellent cytocompatibility of Colloidose^®^ but also its ability to support effective cell spreading, proliferation, and functional integration on the gel surface—features that are essential for wound closure and subsequent tissue remodeling.

**Figure 4.**
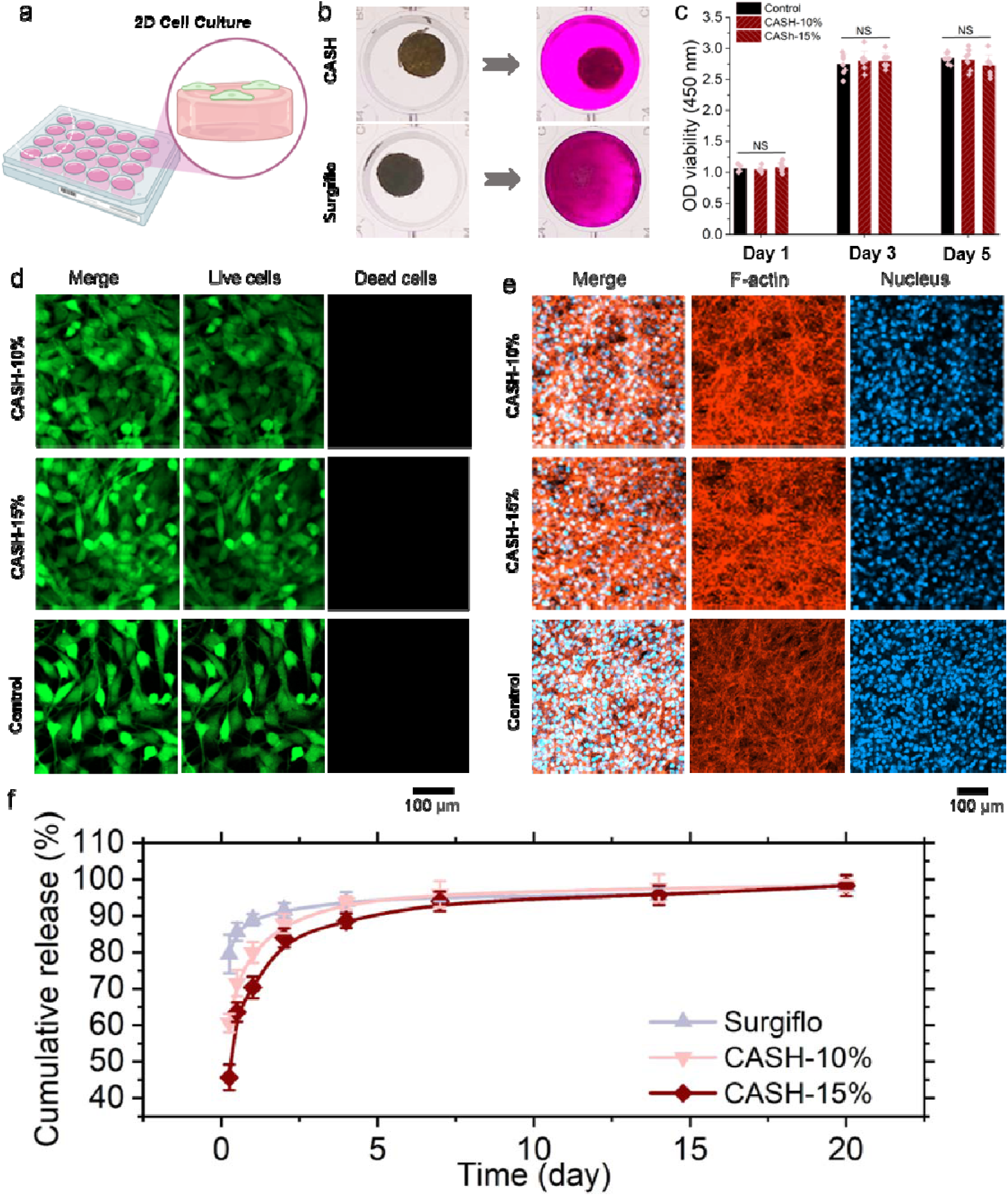
Structural stability, cytocompatibility, and drug release performance of CASH gels. (a) Photograph showing hydrogel immersed in cell culture medium (DMEM containing 10% fetal bovine serum and 1% double antibody at 37 degrees Celsius for 30 minutes), while Surgiflo experienced rapid disintegration and loss of cohesion. (c) CCK-8 assay quantifying fibroblast-like MC3T3 cell viability after 1, 3, and 5 days of culture on top of CASH gels, demonstrating sustained proliferation with no significant cytotoxicity at either concentration (10% and 15% w/v). (d) Live/dead fluorescence staining of MC3T3 cells at Day 3, showing widespread viable cells (green, calcein-AM) and negligible dead cells (red, EthD-1), confirming the cytocompatibility of CASH gels (scale bar: 100 μm). (e) Immunofluorescence staining of α-actinin (red) in MC3T3 cells after 3 days of culture, illustrating well-organized cytoskeletal networks and robust cell spreading on CASH gels (scale bar: 100 μm). (f) Cumulative release profiles of ciprofloxacin, used as a model small-molecule drug, from CASH gels compared with Surgiflo™, demonstrating controlled and sustained release over 21 days with higher drug retention in CASH gels.

Beyond serving as a cell-supportive scaffold, CASH gels were further evaluated as a sustained drug delivery platform. Ciprofloxacin, a model antibiotic, was incorporated into both CASH and Surgiflo™ during preparation, followed by quantitative release profiling. Surgiflo™ exhibited nearly complete drug release within seven days, whereas the CASH formulations displayed markedly prolonged release, with detectable elution persisting for up to 21 days (Figure 4f). This extended release profile suggests that the nanostructured colloidal network effectively retards drug diffusion and thereby prolongs release duration compared to granular systems. This difference is likely attributable to the distinct microstructural architectures of the two matrices. In CASH gels, the continuous nanoscale network provides a high specific surface area and extensive interfacial interactions, which facilitate molecular entrapment and controlled diffusion. In contrast, the granular structure of Surgiflo™, consisting of loosely packed microparticles, lacks cohesive confinement and permits rapid drug escape through interstitial voids. Together, these findings underscore the dual functionality of CASH gels as both a mechanically stable scaffold and a long-acting drug carrier, offering added therapeutic value for post-surgical wound management.

### 2.5. CASH gels induce rapid hemostasis while exhibiting biodegradability and low immunogenicity in rat internal organ hemorrhage models

To quantitatively assess the in vivo hemostatic efficacy of CASH gels, we evaluated both time-to-hemostasis and total blood loss using a standardized rat hepatic hemorrhage model. A parenchymal incision (10 mm in length × 1.5 mm in depth) was surgically created on the hepatic lobe, followed by topical applications using 0.1 mL CASH-10%, CASH-15%, Surgiflo™, or SHAM control (untreated) (Figure 5a). Hemostasis was monitored up to 3 minutes, and blood loss was quantified gravimetrically using pre-weighed filter paper to collect and measure absorbed blood.

**Figure 5.**
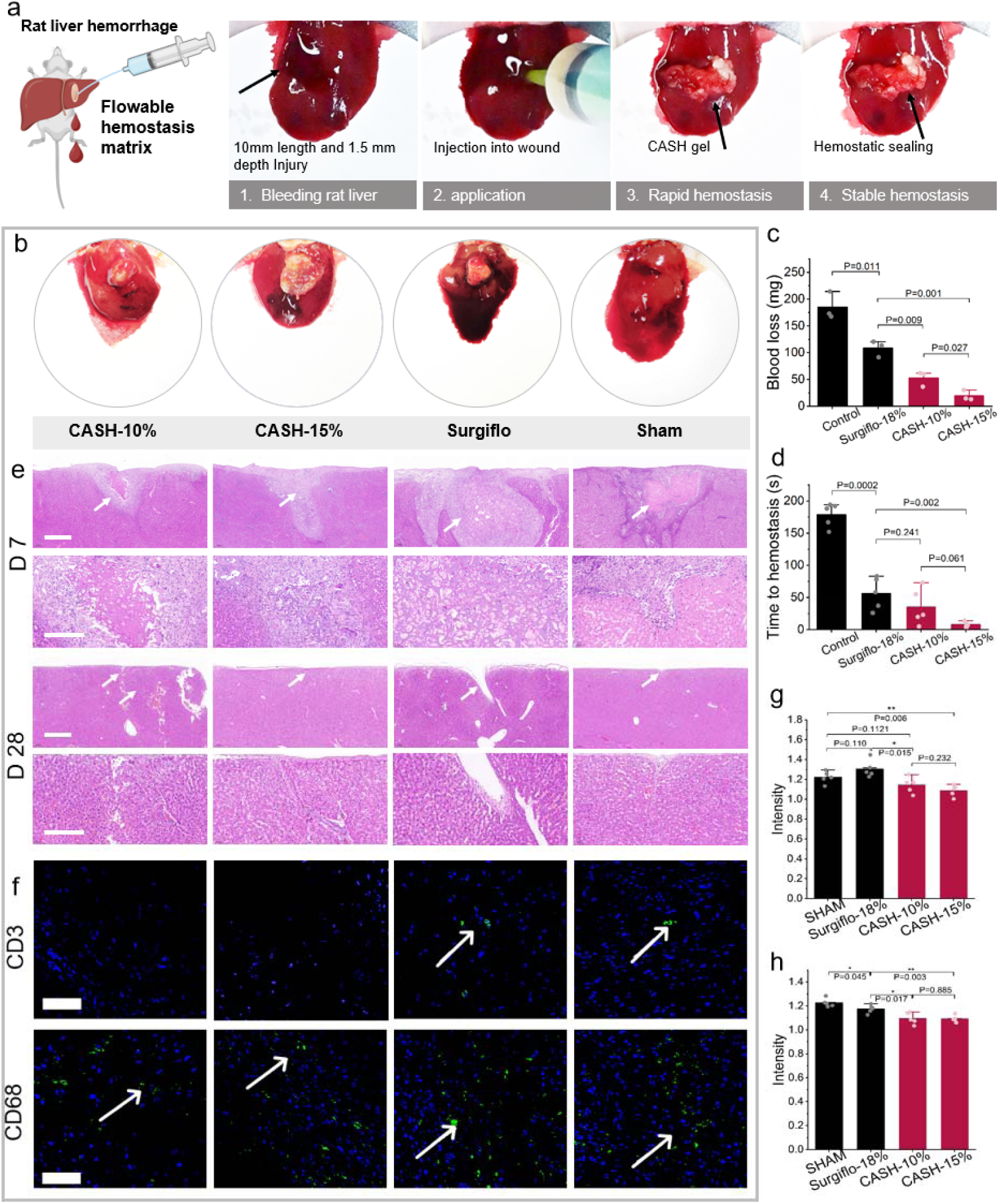
In vivo hemostatic efficacy and biocompatibility of CASH gels in a rat liver injury model. (a) Schematic workflow illustrating the rat hepatic hemorrhage procedure: a standardized 10 mm × 1.5 mm incision was introduced, followed by injection of the flowable matrix, rapid hemostasis, and sealing of the wound. (b) Representative gross images of rat livers after hemostatic treatment with CASH gels (10% and 15% w/v), Surgiflo™, or sham control, demonstrating markedly reduced bleeding in the CASH groups. (c) Quantitative analysis of blood loss and (d) time to hemostasis across treatment groups, showing significantly accelerated hemorrhage control and reduced blood loss in animals treated with CASH gels compared with Surgiflo™ and untreated controls. (e) Representative H&E-stained liver sections at 1 and 4 weeks post-treatment. CASH-treated groups exhibited intact tissue morphology, minimal necrosis, and progressive tissue regeneration, whereas Surgiflo™ and sham groups displayed more pronounced structural disruption. Scale bars = 200 μm. (f) Immunofluorescence staining of liver sections at 1 week post-treatment. DAPI (blue) stains nuclei, while CD3 (green) and CD68 (green) mark infiltrating T cells and macrophages, respectively. Arrows indicate localized immune cell infiltration at the wound site. Scale bars = 100 μm. (g) Normalized fluorescence intensity analysis of CD3 expression, showing comparable or reduced T-cell infiltration in CASH groups relative to controls. (h) Normalized fluorescence intensity of CD68 expression, confirming minimal macrophage-mediated inflammation in the CASH groups.

Compared to the SHAM group (184.96 ± 25.53 mg), both Surgiflo™ and CASH-10% and CASH-15% significantly reduced blood loss, with mean values of 108.37 ± 15.23 mg, 52.89 ± 14.14 mg, and 10.27 ± 9.56 mg, respectively (Figure 5c). Notably, both CASH groups resulted in significantly lower blood loss than Surgiflo™, with the CASH-15% demonstrating the greatest efficacy. Hemostatic kinetics further supported these findings. The SHAM group required 178.18 ± 18.35 s to achieve spontaneous hemostasis via endogenous coagulation, while Surgiflo™ reduced this time to 56.40 ± 24.88 sec. Remarkably, CASH-15% achieved hemostasis in only 7.80 ± 4.32 sec (Figure 5d). Collectively, these results demonstrate that CASH provides markedly superior hemostatic performance, achieving both significantly reduced blood loss and faster hemostasis compared with the clinically established Surgiflo™ matrix. The combination of rapid mechanical sealing and effective clot stabilization likely underpins this enhanced in vivo efficacy.

Further evaluation of the biocompatibility of the gelatin-based hemostatic matrices was performed through histological and immunofluorescence analyses of rat liver tissues, aiming to assess wound healing progression and local inflammatory responses. Hematoxylin–eosin (H&E) staining on postoperative days 7 and 28 demonstrated ongoing tissue regeneration with generally low levels of inflammation across all treated groups, including Surgiflo™ and CASH(Figure 5e). At day 7 post-implantation, residual Surgiflo™ material was still detectable at the wound site, encapsulated by newly formed connective tissue. This finding suggests a moderate foreign body response with gradual matrix degradation. By day 28, the Surgiflo™ matrix was almost completely resorbed, accompanied by new tissue integration and visibly reduced inflammation, reflecting a favorable biocompatibility profile. In the CASH-treated groups, only trace amounts of residual gel were observed as early as day 7, indicating a faster degradation profile. Histological sections revealed abundant fibroblast infiltration, active extracellular matrix remodeling, and limited presence of granulocytes and macrophages. These features are indicative of an active yet well-regulated healing process, characterized by effective tissue reconstruction with minimal prolonged inflammation. By contrast, the untreated SHAM group showed only superficial wound closure by day 28. The underlying tissue remained disorganized and infiltrated with inflammatory cells, suggesting incomplete resolution of injury and suboptimal healing outcomes.

To further characterize the immune response, immunofluorescence staining was performed at postoperative day 7. CD3+ T-cell infiltration, a marker for adaptive immune activation, was significantly higher in the SHAM and Surgiflo™ groups compared to the CASH groups. However, the immune cell infiltration in the Surgiflo™ group was still markedly lower than in the SHAM group, indicating a moderated host response. Both CASH-10% and -15% showed minimal CD3+ infiltration, with no significant difference between concentrations (Figures 5f, g), suggesting reduced T-cell-mediated inflammatory signaling. Similarly, CD68+ macrophage and foreign body giant cell densities were highest in the SHAM group, followed by Surgiflo™, and lowest in CASH groups. The differences between Surgiflo™ and CASH gels were not statistically significant (Figure 5h). While Surgiflo™ demonstrates favorable biocompatibility and moderate immune response suitable for clinical use, CASH gels appears to further enhance the healing environment by promoting rapid degradation and minimizing local inflammation. These findings suggest that CASH gels may serve as a promising alternative with improved immunomodulatory potential.

### 2.6 Colloidose^®^ Exhibits Effective Hemostatic Performance in Large-Animal Models

Building on the remarkable hemostatic efficacy of CASH gels demonstrated in both in vitro assays and rat liver hemorrhage models, we advanced the platform into a Class III medical device, Colloidose^®^, and conducted systematic evaluations in large-animal preclinical studies. The standard Colloidose^®^ formulation uses 10% (w/v) CASH gel, chosen for its balance of injectability, strength, and ease of use in surgery. To assess its translational potential, further experiments were carried out in Beagle dog and porcine models under various bleeding scenarios. We initially assessed the hemostatic performance of Colloidose^®^ using an ex vivo porcine liver laceration model designed to replicate clinically significant parenchymal hemorrhage. A standardized deep linear incision (3.0 cm × 0.5 cm × 1.0 cm) was made to induce uncontrolled bleeding. After a 2-second free-bleeding interval to simulate initial blood loss, either Colloidose^®^ or Surgiflo™ was applied according to the respective manufacturer’s instructions. Both materials achieved effective hemostasis. Quantitative measurements showed that Colloidose^®^ reached hemostasis within 87.5 ± 34.2 seconds, while Surgiflo™ required 134.5 ± 100.6 seconds, and the untreated group required 256.9 ± 42.1 seconds (Figure 6b). These results indicate that both Colloidose^®^ and Surgiflo™ are effective in achieving hemostasis under clinically relevant bleeding conditions. To further evaluate the biosafety of Colloidose^®^, histopathological examinations were conducted on major organs, including the heart, liver, spleen, lungs, and kidneys, on postoperative days 7 and 28. No signs of inflammation, necrosis, or histopathological abnormalities were observed (Figures S3 to S5). In addition, complete blood count (CBC) analysis showed that hematological parameters, including white blood cell, red blood cell, and platelet counts, remained within physiological ranges in all treated animals (Figures S6 to S8), supporting the absence of systemic toxicity.

**Figure 6.**
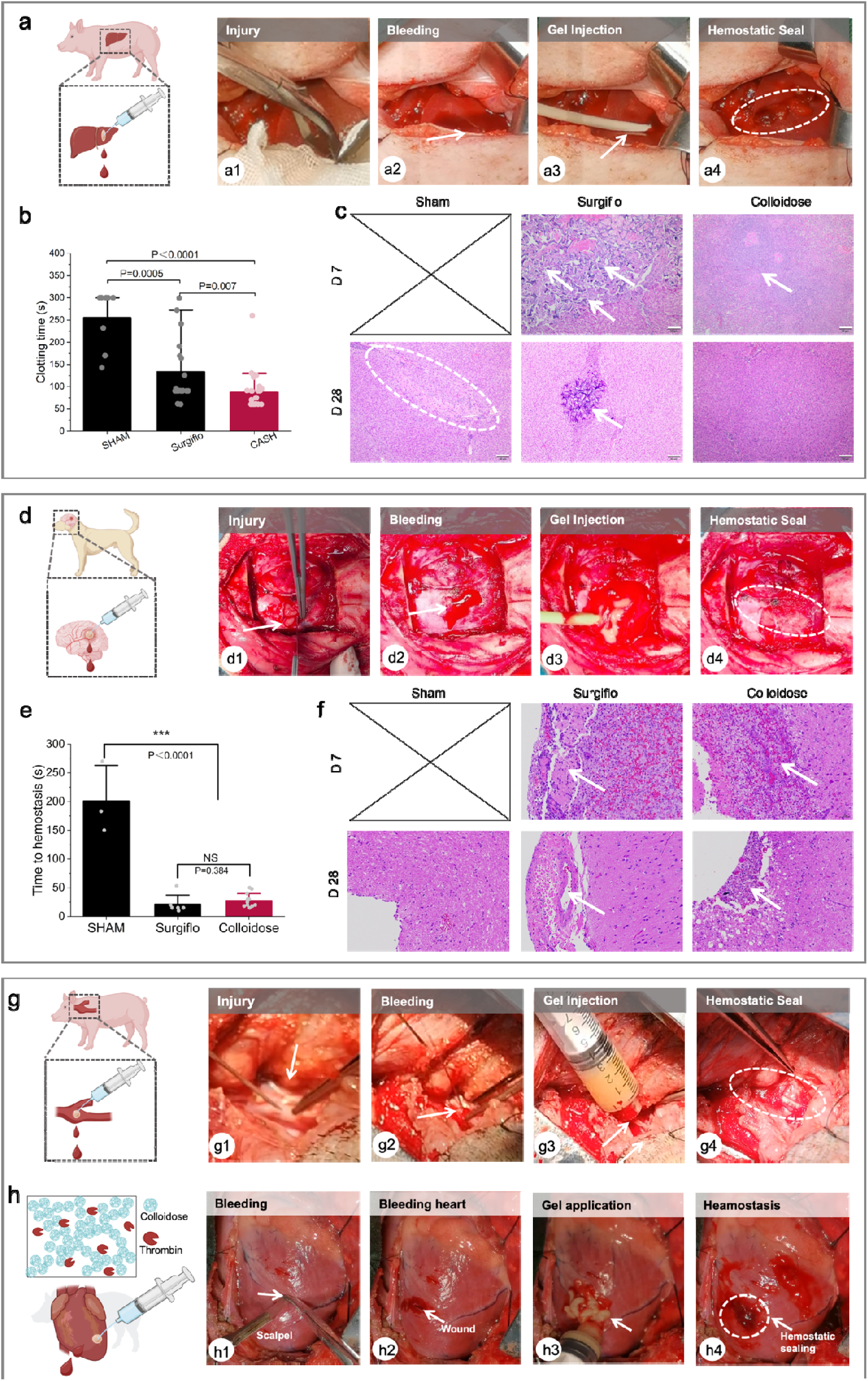
Hemostatic performance of Colloidose^®^ in preclinical large-animal models. (a) In vivo porcine liver hemorrhage model. Sequential images show incision, active bleeding, Colloidose^®^ injection, and stable hemostatic sealing. (b) Quantitative analysis of time to hemostasis in porcine liver injuries, demonstrating significantly shorter hemostasis with Colloidose^®^ compared to Surgiflo™ and SHAM controls. (c) Representative H&E-stained sections of ex vivo porcine liver wounds post-treatment. Colloidose^®^-treated samples showed rapid integration with host tissue and minimal residual material, whereas Surgiflo™ retained particulate remnants and SHAM controls displayed extensive hemorrhagic regions. (d) Beagle dog cerebral hemorrhage model. Sequential intraoperative images illustrate Colloidose^®^ injection at the cortical wound site and subsequent hemostatic sealing. (e) Hemostasis times for canine brain injuries. Colloidose^®^ achieved significantly faster bleeding control compared with Surgiflo™ and SHAM groups. (f) Representative H&E-stained sections of ex vivo canine brain tissue, showing well-preserved parenchymal morphology and minimal inflammatory infiltration in the Colloidose^®^ group, in contrast to residual material and localized inflammation in Surgiflo™ and untreated tissues. (g) Colloidose^®^ gels rapidly sealed a standardized poorcine femoral arterial defect under pulsatile flow, preventing further blood loss within 3 min. (h) Injection of thrombin-loaded Colloidose^®^ (100 U/mL) enabled immediate sealing of Porcine myocardial wound, achieving complete hemostasis without rebleeding during observation.

At the implantation site, differences in degradation kinetics were observed between the two hemostatic matrices. Hematoxylin and eosin (H&E)-stained tissue sections on postoperative day 7 showed that Colloidose^®^ had partially integrated into the surrounding host tissue, with only minimal eosinophilic remnants visible, suggesting ongoing biodegradation. In contrast, Surgiflo™ maintained well-defined particulate structures with limited signs of resorption at the same time point (Figure 6c). By day 28, Colloidose^®^ underwent near-complete degradation, which was associated with organized collagen matrix deposition and minimal residual inflammatory cell infiltration. In comparison, residual Surgiflo™ fragments remained present and were partially surrounded by multinucleated foreign body granulomas, indicating a slower resorption process. These observations suggest that Colloidose^®^ facilitates rapid material clearance and supports a local environment conducive to regenerative wound healing through its biodegradability and integration with host tissue.

Given the clinical urgency for minimally invasive local hemostatic interventions in cases of cerebral hemorrhage, we further evaluated the efficacy of Colloidose^®^ using a canine brain injury model. A standardized linear cortical wound measuring 1.0 centimeter in length was surgically induced in Beagle dogs, followed by minimally invasive administration of Colloidose® through direct intracerebral injection (Figure 6d). Hemostatic performance was quantitatively assessed and compared among Colloidose^®^, Surgiflo™, and untreated controls.

Colloidose® achieved rapid hemorrhage control, with a mean time to hemostasis of 39.8 ± 15.9 sec. This was significantly shorter than that of the untreated group (203.3 ± 32.2 sec) and also numerically lower than that of Surgiflo™ (51.7 ± 26.0 sec), although the difference between Colloidose^®^ and Surgiflo™ did not reach statistical significance (Figure 6e). These findings indicate that both Colloidose^®^ and Surgiflo™ effectively reduce bleeding time in this neural injury model compared to untreated controls. The lower mean time to hemostasis observed with Colloidose^®^ suggests a potential performance advantage, warranting further investigation. Additionally, the result represents an improvement over its performance in hepatic hemorrhage models, demonstrating the robust and consistent hemostatic activity of Colloidose^®^ across distinct anatomical and physiological contexts, including highly vascular and delicate neural tissues.

Histological analysis using hematoxylin and eosin (H&E) staining provided insight into material degradation and local tissue responses (Figure 6f). At one week post-implantation, residual material from both Colloidose^®^ and Surgiflo™ remained detectable at the injury site, suggesting comparable early-stage biodegradation kinetics. Mild inflammatory infiltration, primarily consisting of lymphocytes and macrophages, was observed at the material–tissue interface in both treatment groups, consistent with a typical acute-phase response. By postoperative week six, both materials had undergone near-complete degradation, with only minimal residual fragments detected within the brain parenchyma. Inflammatory responses were substantially reduced and largely confined to perivascular regions, with no histological evidence of chronic granulomatous reactions or tissue necrosis. These findings indicate that both Colloidose^®^ and Surgiflo™ demonstrate favorable degradation profiles and biocompatibility in neural tissue. Furthermore, Colloidose^®^ was able to achieve effective and timely hemostasis in cerebral applications while exhibiting degradation and inflammatory characteristics similar to those of clinically validated materials. These properties support its potential suitability for neurosurgical use, where both precision and biocompatibility are essential.

Following the demonstrated effectiveness of Colloidose^®^ in conventional local hemostatic applications, its performance was further investigated under more challenging arterial bleeding conditions. The initial assessment utilized a porcine femoral artery injury model, in which a standardized circular defect (3 mm in diameter) was surgically introduced to generate pulsatile hemorrhage (Figure 6g). Colloidose^®^ was immediately applied to the defect and rapidly sealed the arterial breach, likely due to its substantial mechanical strength. No additional bleeding was observed within three minutes of application, indicating successful hemostasis under high-pressure vascular conditions. In view of the established clinical practice of combining flowable hemostatic matrices with thrombin, we next evaluated the performance of the current Colloidose^®^ co-administered with thrombin in a porcine cardiac injury model. Linear myocardial defects were surgically created, and the Colloidose^®^–thrombin composite (100 U/mL) was delivered directly into the wound (Figure 6h). This formulation achieved rapid hemostasis through an integrated mechanical and biochemical mode of action. Its intrinsic mechanical robustness enabled the material to resist the dynamic hemodynamic forces generated by cardiac contraction, while its high conformability facilitated close adherence to the wound surface. Concurrently, the embedded thrombin catalyzed fibrin polymerization, thereby accelerating the physiological coagulation cascade. As a result of this combined mechanism, bleeding at the wound margins ceased within seconds. A visible color transition from yellow to red was noted, indicating rapid fluid uptake and matrix activation. Complete hemostasis was achieved within three minutes, and no rebleeding events occurred during the postoperative observation period. These findings suggest that the Colloidose^®^–thrombin composite may serve as an effective adjunct in cardiac surgical applications requiring prompt and localized hemorrhage control.

### 2.7 Clinical trials confirm outstanding hemostatic efficacy of Colloidose^®^ in multiple surgical fields

We conducted a multicenter, single-blind, randomized clinical study comparing the hemostatic efficacy of Colloidose^®^ and Surgiflo™ across three surgical departments: Orthopedics, Gynecology, and General Surgery (Figure 7a). A total of 348 patients were enrolled and randomly assigned in a 1:1 ratio to receive either Colloidose^®^ (test group) or Surgiflo™ (control group). These departments were selected to reflect a broad range of clinical hemostatic challenges. In Orthopedics, spinal surgeries require materials with high injectability and minimal swelling due to proximity to neural structures. General Surgery and Gynecology, dominated by laparoscopic procedures, demand compatibility with minimally invasive delivery systems. Demographic characteristics of all patients are presented in Table S1. Eligible participants exhibited intraoperative capillary, venous, or small arterial bleeding that was refractory to conventional techniques such as electrocautery or compression, necessitating adjunctive hemostatic intervention.

**Figure 7.**
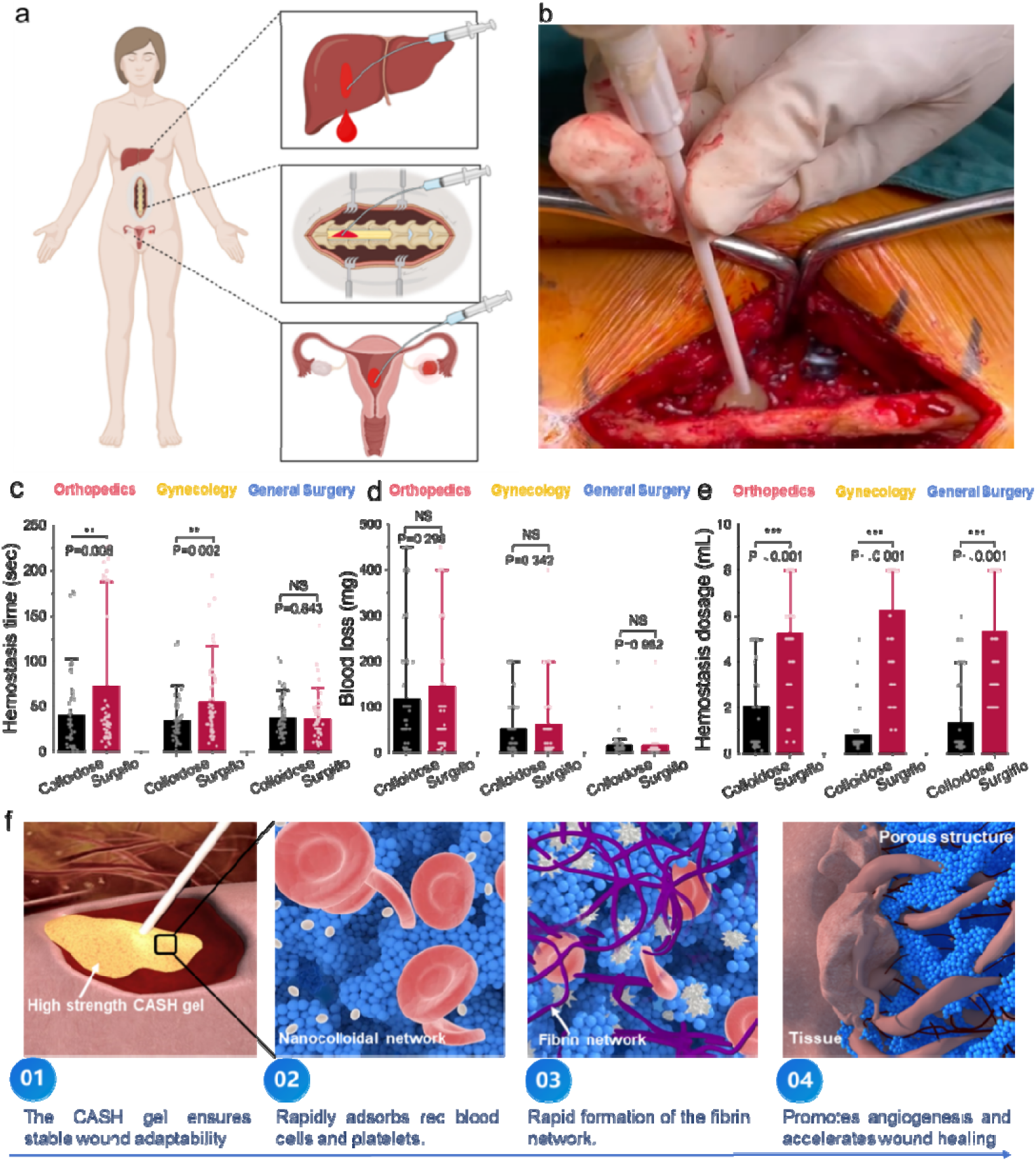
Clinical evaluation of Colloidose^®^ in managing persistent surgical bleeding and proposed hemostatic mechanism. (a) Schematic representation of the multicenter, randomized clinical study design evaluating CASH gels in three surgical specialties: orthopedics, gynecology, and general surgery. (b) Intraoperative photograph demonstrating direct application of CASH gel achieving rapid hemostasis during spinal surgery. (c) Quantitative comparison of hemostasis times in orthopedic, gynecological, and general surgery procedures, showing that Colloidose^®^ significantly shortened bleeding control times compared with Surgiflo™. (d) Quantitative analysis of intraoperative blood loss across all surgical categories, demonstrating lower volumes in the Colloidose^®^ groups for orthopedic and general surgery cases, and comparable performance between Colloidose^®^ and Surgiflo™ in gynecology. (e) Quantification of effective hemostatic dose, showing that Colloidose^®^ required markedly smaller application volumes than Surgiflo™, highlighting superior efficiency in clinical use. (f) Schematic illustration of the multimodal hemostatic mechanism of Colloidose^®^, highlighting strong wound sealing, rapid adsorption of red blood cells and platelets, accelerated fibrin network formation, and porous architecture that facilitates angiogenesis and tissue regeneration.

Colloidose^®^ demonstrated shorter mean hemostasis times across all departments compared to Surgiflo™ (Orthopedics: 168.0 ± 65.7 sec vs. 180.0 ± 112.2 sec; General Surgery: 168.0 ± 65.7 sec vs. 180.0 ± 112.2 sec; Gynecology: 168.0 ± 65.7 sec vs. 180.0 ± 112.2 sec) (Figure 7c). Additionally, the 3-minute hemostasis rate was slightly higher for Colloidose® (98.95%) than for Surgiflo™ (94.89%), although both groups achieved 100% by 5 minutes (Table S2). In terms of intraoperative blood loss, Colloidose^®^ was associated with lower mean volumes in Orthopedics (116.5 ± 124.3 mg vs. 144.9 ± 162.2 mg) and General Surgery (49.2 ± 56.2 mg vs. 61.2 ± 76.4 mg) compared to Surgiflo™. Notably, in Gynecological procedures, both materials demonstrated comparable performance, with nearly identical mean blood loss values (Colloidose^®^: 16.6 ± 28.5 mg; Surgiflo™: 16.4 ± 29.3 mg), highlighting the effectiveness of both agents in this context. Colloidose^®^ required significantly lower application volumes across all surgical types (Orthopedics: 2.1 ± 1.9 mL; Gynecology: 0.81 ± 0.95 mL; General Surgery: 1.3 ± 1.6 mL) compared to Surgiflo™ (5.2 ± 2.7 mL, 6.2 ± 2.5 mL, and 5.4 ± 2.5 mL, respectively). This approximately 3:1 reduction in volume required to achieve hemostasis supports Colloidose^®^’s efficient wound sealing capacity and may contribute to both improved procedural safety and reduced treatment costs. Overall, these clinical findings support Colloidose^®^ as a reliable adjunct for managing challenging surgical bleeding, particularly where conventional methods prove inadequate. The comparable outcomes in gynecologic procedures also affirm the performance of both products in minimally invasive, capillary bleeding scenarios.

Based on comprehensive preclinical and clinical findings, we propose a mechanistic explanation for the hemostatic efficacy of Colloidose^®^. First, the material provides physical sealing and mechanical stabilization. Upon injection, Colloidose^®^ rapidly reconstructs into a cohesive colloidal network that conforms to irregular wound geometries. Its intrinsic self-healing ability, together with high mechanical integrity and resistance to dispersion, ensures robust and sustained compression of the bleeding site, even under high-pressure or dynamic conditions such as tissue movement or continuous blood flow. Second, the material supports the biological process of hemostasis. The colloidal network structure passively concentrates blood components including erythrocytes and platelets, while simultaneously adsorbing fibrinogen to accelerate fibrin polymerization. In addition, the matrix may sequester endogenous growth factors, thereby creating a favorable microenvironment that promote tissue regeneration. Through the combination of mechanical stabilization and biochemical support, Colloidose^®^ achieves rapid and effective hemostasis while facilitating subsequent wound healing.

## **3.** Conclusion

In this study, we developed CASH gels (commercially named Colloidose^®^) as a second-generation flowable hemostatic matrix and demonstrated their successful translation from preclinical research to clinical application. Comparing with more conventional design of flowable gelatin matrix using hundreds micrometer-sized gelatin granules, the reversibly crosslinked colloidal network endows Colloidose^®^ with superior mechanical strength, rapid self-healing capability, and favorable injectability, enabling stable defect-conforming sealing even under high-pressure bleeding conditions. Systematic characterization and comparative analyses against commercial granular gelatin hemostatic matrices confirmed the advances of Colloidose^®^ matrix to achieve faster hemostasis, reduce blood loss, and minimize required dosage. Their biodegradability and low immunogenicity further support safe in vivo application. Extensive validation in small- and large-animal hemorrhage models confirmed robust efficacy across diverse bleeding scenarios, while multicenter clinical trials in orthopedics, gynecology, and general surgery demonstrated significant advantages of Colloidose^®^ over currently available commercial comparators, including shorter hemostasis times and lower application volumes. These findings highlight Colloidose^®^ as a reliable adjunct for managing surgical bleeding, particularly in anatomically complex or compression-intolerant sites where existing hemostatic agents show limitations. Overall, this work establishes CASH gels as a clinically translatable self-healing biomaterial that not only advances hemostatic technology but also provides a generalizable strategy for developing injectable and moldable biomedical devices. The integration of mechanical resilience, self-healing, and biological compatibility may inspire broader applications of self-healing colloidal gels towards applications in regenerative medicine, drug delivery, and beyond.

## Data Availability

All data produced are available online

## Acknowledgements

This work was granted by National Key Research and Development Program of China (No. 2022YFC2403002), National Natural Science Foundation of China (No.52403152), Natural Science Foundation of Liaoning Province (Grant No. 2024-BSBA-02), Medical-Engineering Interdisciplinary Joint Fund of Dalian University of Technology (Grant No. DUT24YG102).

## Ethics Statement

All clinical procedures in this study were conducted in accordance with the ethical standards of the institutional and/or national research committee and with the 1964 Helsinki Declaration and its later amendments. The clinical protocol was reviewed and approved by the Institutional Review Board (IRB) of West China Hospital, Sichuan University (Approval No. HX-IRB-AF-19-V4.0). Written informed consent was obtained from all participants prior to their inclusion in the study. All personal data were anonymized to ensure patient confidentiality.

