## Supplementary Materials for "Bench-to-bedside translation of Self-Healing colloidal hydrogels as 2^nd^ generation design of Flowable Hemostatic Matrix: From Preclinical evaluation to Human Clinical Trials"

### 1. Materials and Methods

#### 1.1 Materials

CASH was supplied by Shenzhen Huanova Biotechnology Co., Ltd.(Batch number: 20200302, specification: 0.5g/branch); 0.5 g of CASH was placed in a 10 ml syringe and saline was injected in another syringe to achieve a 10%, 15% and 17.8% volume fraction(marked as CASH-10% and CASH-15% and CASH-high). The saline was injected into the CASH by connecting the two syringes using luer connector and pushing back and forth to mix them thoroughly. The air in the syringe is removed to obtain an injectable colloidal gel. Srugiflo^TM^, flowable hemostatic matrix without thrombin (Jonson & Jonson Medical Devices companies, Batch No.: 255422; specification: 8 mL/bottle).

To quantify the mass fraction of the hemostatic matrix, we prepared the hemostatic fluid according to standard methods. Mix Surgiflo with 2 ml of physiological saline, repeatedly squeeze the syringe 10 times, take 1 ml of the completely mixed Surgiflo, weigh it, and record the wet weight m1. Then freeze-dry it, weigh it again, and record the dry weight m2, mass fraction= m2/ m1,the mass fraction is about 18% at this time, denoted as Surgiflo. To fairly compare with CASH at the same mass fraction, we further obtained Surgiflo with mass fractions of 10% and 15% by adding saline, and denoted as Surgiflo-low and Surgiflo-medium respectively）

#### 1.2 Characterizations

CASH and Surgiflo were directly subjected to scanning electron microscope (SEM, Nova Nano SEM 450) observation after freeze-drying to obtain their morphological characteristics. CASH and Surgiflo were respectively co-incubated with red blood cells and buffy coat for 1 hour, then fixed with polyformaldehyde, dehydrated with different concentrations of ethanol-water and ethanol, then the micro-morphology was observed by SEM after spray-gold treatment.

Fourier transform infrared spectroscopy (FT-IR)（ThermoFisher Nicolet iS5）was used to analyze the chemical composition of CASH and Surgiflo. After freeze-drying, CASH and Surgiflo were ground to a fine powder, and compacted into disks. The scanning speed was set to 0.2 cm/s, 32 scans were carried out with a resolution of 4 cm^-1^, and 10 times were performed for each detection.

#### 1.3 Mechanical Characterization

#### The rheology properties were performed by Discovery hybrid rheometer (DHR, TA instrument) with parallel plate (diameter 20 mm). Temperature is set to 25 °C, and the operating gap is 1000μm. The shear-thinning behavior was determined by measuring their viscosity and changing with shear rate from1 s^-1^ to 100 s^-1^. After a tiem sweep with a strain of 0.5% and frequency of 1 rad/s for 4 mins, the amplitude sweep time sweep is carried out with 1 rad/s frequency and strain from 0.5 to 100 %, then continue with the time sweep described above to perform the self-healing properties of the samples, all samples were tested by three cycles between continuously destroyed and recovered holding. After each cycle, the recovery rate (%) of the storage modulus (G') of the sample under time sweep relative to the initial modulus before the huge strain was compared to quantify the self-healing ability. The frequency is set from 0.1 rad/s to 100 rad/s and a constant 0.5 % shear strain is set to characterize the viscoelastic properties.

All syringes with CASH and Surgiflo samples were fixed vertically on a homemade scaffold in a compression mode. Use a compression mode of a universal testing machine (E43, MTS instrument, USA) to press the plunger of syringe at a constant speed of 2 mm/min.

#### 1.4 In vitro hemostatic activity determination

A custom-built burst pressure testing system for hemostatic gels was constructed using a pressure sensor and a peristaltic pump. A 5 mm × 5 mm incision was made on pig skin, and the incision was sealed with CASH-10%, CASH-15%, and Surgiflo, respectively. Blood was then injected at a flow rate of 0.1 ml/s until it began to flow out from the sealed incision. The burst pressure during this process was recorded, with each sample tested four times.To assess the in vitro blood absorption capacity of the hemostatic gels, 0.5 ml of CASH and Surgiflo were placed on filter paper, and blood was continuously added to the gels until saturation. The weight was measured after saturation, and the mass of blood absorbed by the filter paper was subtracted to calculate the blood absorption capacity of each sample with different mass fractions. Each sample was tested six times.ROTEM/TEG was used to investigate the in vitro coagulation performance, we mixed the samples with citrate-anticoagulated pig blood (3.2% sodium citrate) at volume ratios of 1:5 and 1:10. After mixing, 300 µL of the sample or blood was combined with 20 µL of 0.2 M/L calcium chloride to reintroduce calcium ions, and an intrinsic pathway activator (Factor XII) was added for testing.

Blood was collected from the ear veins of Japanese rabbits and stored in blood collection tubes containing sodium citrate. Centrifuge at 275G for 10 minutes, and retain the platelet-containing plasma after mixing PRP and PPP. Take 0.2 g of the sample and place it in a centrifuge tube, add 0.5 ml of platelet-containing plasma to immerse the sample, and incubate it in a water bath at 37 °C for 10 minutes. The soaked samples were dehydrated with 20%, 40%, 60%, 80% and 100% ethanol aqueous solution respectively, and then fixed by adding 0.5ml of paraformaldehyde. After freeze-drying, the surface platelet adsorption was observed by SEM.

#### 1.5 Cytotoxicity evaluation of CASH

MC3T3 were used to evaluate the cytocompatibility of the CASH hydrogel. Surgiflo, due to its disintegration in water, was unsuitable for surface cell culture. Dulbecco's Modified Eagle Medium (DMEM) with high glucose, supplemented with 1% penicillin-streptomycin solution and 10% fetal bovine serum (FBS), was used as the complete culture medium. CASH was prepared into 5 mm diameter and 2 mm thick discs and placed in a 96-well plate. MC3T3 were initially seeded into the 96-well plate containing 500 µL of the complete medium. The cells were cultured for 1, 3, and 5 days at 37°C in a humidified incubator with 5% CO₂. Cell viability was assessed using the CCK-8 assay. All experiments were performed in six replicates, and the average values were calculated. Cells incubated in complete medium without any hydrogel were used as the control group.The viability of MC3T3 on CASH discs was measured using the LIVE/DEAD® Viability/Cytotoxicity Kit (Invitrogen, Shanghai, China), following the manufacturer's protocol. Specifically, 2 µM calcein-AM and ethidium homodimer-1 were added to the culture plate and incubated for 10 minutes at 37°C. The hydrogel discs were then washed three times with the culture medium, and cell viability was observed using a laser confocal microscope (Zeiss LSM980, Germany). To observe the morphology of MC3T3, Alexa Fluor 594 Phalloidin (Invitrogen, USA) and 4',6-diamidino-2-phenylindole (DAPI) (Solarbio, China) were used to stain F-actin and cell nuclei, respectively. Spheroid samples of different sizes were collected on days 1 and 7 and fixed overnight with 4% paraformaldehyde (PFA). The samples were permeabilized with 0.1% Triton X-100 (Solarbio, China) for 20 minutes, blocked with 1% bovine serum albumin (BSA) (Solarbio, China) for 45 minutes, and washed with PBS. The fixed samples were then stained with Alexa Fluor 594 Phalloidin for 45 minutes, followed by DAPI staining for 10 minutes, and washed three times with PBS. Fluorescent images were examined using a laser confocal microscope (Zeiss LSM980, Germany).

The drug release kinetics of Ciprofloxacin loaded on Colloidaose-10%, CASH-15% and Surgiflo were determined in a phosphate buffer at a pH of 7.4. 10 mg of the drug was loaded by 0.5 g matrix through mixing with the saline. Matrix was encapsulated in a semi-permeable membrane (3500Da), then dissolved in phosphate buffer (50 mL) with pH 7.4 The drug release kinetics were studied by recording (triplicate) absorbance measurements at λ_max_ = 272 nm using UV–Vis spectrophotometer up to 20 days.

#### 1.6 Animal experiment

The in vivo hemostatic properties of matrix were assessed by using mouse liver bleeding models. All animal experiments were approved by the Institutional Review Board of Dalian University of technology. the in vivo hemostatic performance of the CASH and Surgiflo was evaluated using a mouse liver puncture bleeding model (SD mice, 8weeks g, male). The mice were anesthetized with isoflurane and then secured onto a surgical corkboard. The liver was exposed through an abdominal incision, and gauze was used to remove tissue fluid surrounding the liver. Pre-weighed filter paper was placed directly beneath the liver and separated from the abdominal incision with plastic wrap. Prepare a 10mm long and 1.5mm deep wound on the surface of rat liver using a willow blade and the Collodiose or Surgiflo was immediately put onto the wound. The SHAM group did not receive any hemostatic treatment until the blood coagulated After the bleeding stopped, the weight of the absorbed blood was weighed and the bleeding time was recorded and compared to SHAM group. Each group contained 5 mice. The regenerated wound tissues at various healing stages were examined using hematoxylin-eosin (H&E) staining on day 7 and day 28. At the same time, the inflammation status of the wound area was evaluated using immune fluorescence staining of CD3 and CD68. Instruments and materials: The Monitor ( iPM12 Vet ) ; Microinjection pump ( WZS-50F2 ) ; Electric attractor (YB-DX23D) ; High frequency surgical scalpel (GD350-D) ; One-time use of tracheal intubation ( Zhejiang Haisheng, 5.0) ; Medical gauze piece (Shaoxing Zhende, 7.5cm *7.5cm-8P, 5 pieces / bag ). Isoflurane (Ringpu in Tianjin , 100 mL/bottle); Scopolamine injection (Jilin Huamu, 0.3 mg/mL).

Bama minipigs with 5 males and 16 females (weighing 20-30 kg, 6-12 months) were provided by Wujiang Tianyu Biological technology Co., Ltd. Anesthesia ventilator ( WATO EX-20VET ) ; A total of 21 Bama minipigs（5males and 16 female）were purchased from Wujiang Tianyu Biological technology Co., Ltd Anesthesia ventilator (WATO EX20VET). The 12h light/dark cycle was applied to these minipigs with no limit of diet, and temperature was maintained at 25 degrees Celsius.

All animal experiments in this study were designed and approved by the Biological and Medical Ethics Committee of the Dalian University of Technology. Twenty-one Bama minipigs were divided into three groups, including 3 in the SHAM group, 6 in the Surgiflo, and 12 in the CASH. Scopolamine (0.01 mg/kg) was injected into minipigs to reduce respiratory secretions. The minipigs were anesthetized by isoflurane, and vein channels were established. After the surgical site was disinfected, the incision with a length of 3.0 cm and a depth of 0.5 cm was performed in the liver. The wound site of the minipigs in the SHAM group was not treated but pressed to stop bleeding after 5 minutes. After 2 s of bleeding at the incision of animals in the Surgiflo group and CASH group, the blood on the surface was gently wiped off with sterile gauze, and the hemostatic gel was squeezed on the bleeding position. The bleeding was observed every 20 s until complete hemostasis. The product and tissue adhesion were subjectively evaluated. Within 3 days after surgery, 2.0 g cefazolin sodium was intramuscularly injected, twice a day. A series of behavioral phenomena such as appearance signs, behavioral activities, body temperature, local irritation, gland secretion, fecal traits, and food intake were observed regularly after the operation.

Blood biochemistry and coagulation were collected before and after the operations, and the results were statistically analyzed and compared. After the experimental animals were sacrificed, the anatomical process should be carefully observed whether there are allergies, infections, hematomas, coagulation disorders, adhesions, and other complications.

One minipig should be randomly selected at 4 weeks after the operation for pathological analysis of the main organs (including liver, kidney, heart, lung, spleen, brain) and all visible lesions. A frozen sectioning procedure was conducted on each liver incision block, all tissue samples were processed by H&E staining and then imaged using a stereoscope microscope. As shown in table 1, we Set 3 observation time points: 1 week, 2 weeks and 4 weeks after operation.

Table.1 Number of animals at the different observation point

| Groups | Time | | |
| --- | --- | --- | --- |
|  | 1 week | 2 weeks | 4 weeks |
| Sham-operated | N/A | N/A | 3 |
| Surgiflo | 3 | N/A | 3 |
| CASH | 3 | 3 | 6 |

(Plus: N/A: Not Applicable, 'not applicable 'means' this column (for this group) is not applicable')

#### 1.11 Statistical analysis

Statistical analyses were conducted using One-way ANOVA with GraphPad Prism 8 (Graphpad Software, USA), error bars represent standard deviation, and significance was analysed using the t-test, the differences are significant at p < 0.05 marked as *, marked as ** at p<0.01, and marked as *** at p<0.001.

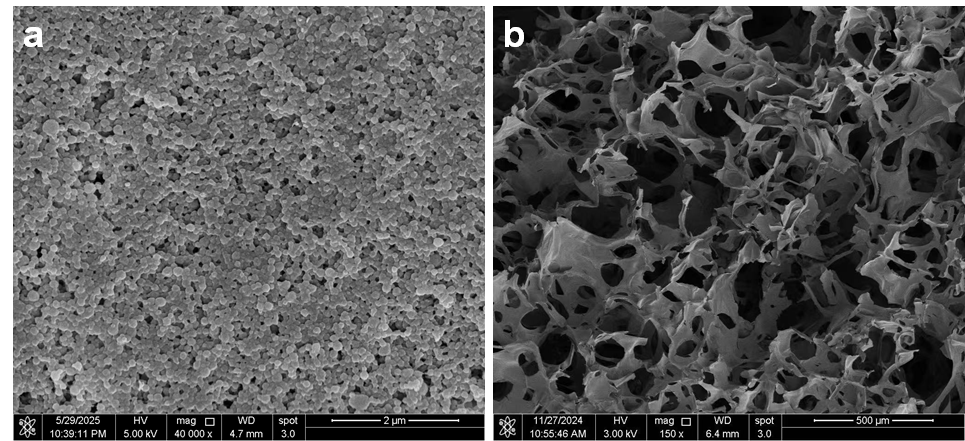

Figure S1. SEM images of Colloidose^®^ and Surgiflo™ flowable matrix after gel preparation. Representative microscopic SEM images highlighted the distinct network morphologies of the two hemostatic matrices.

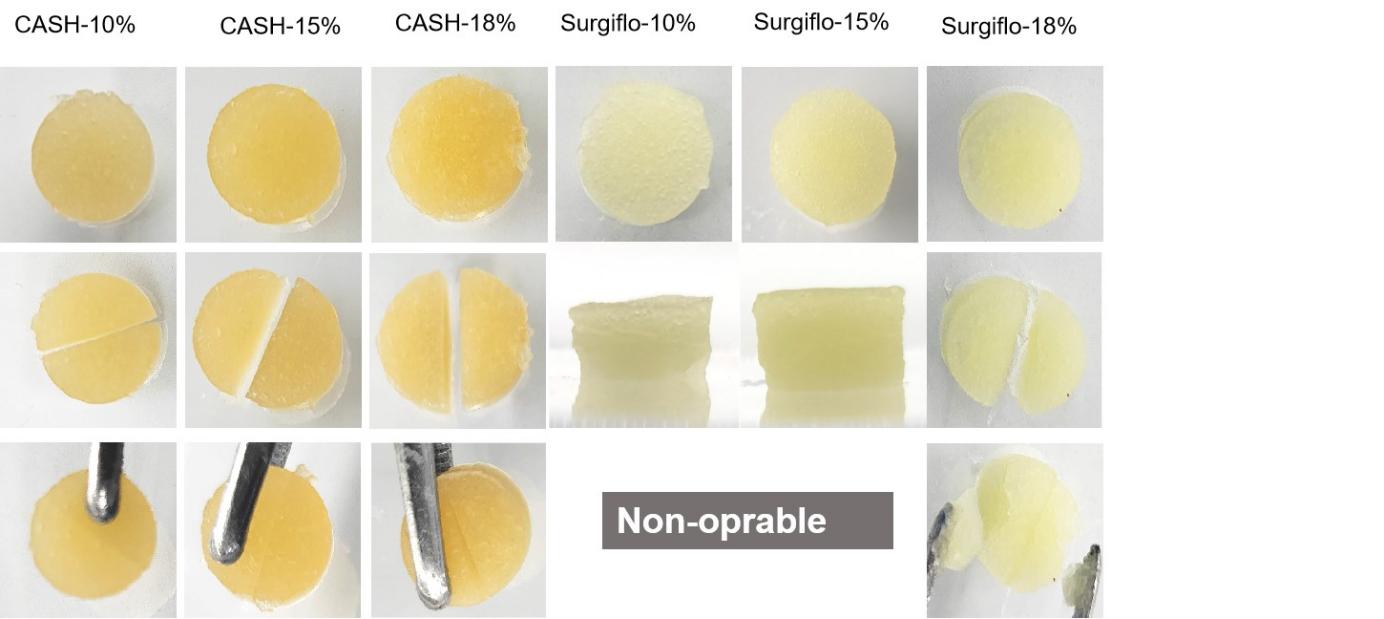

Figure S2.The self-healing performance of Colloidose^®^ and Surgiflo™ flowable matrix.

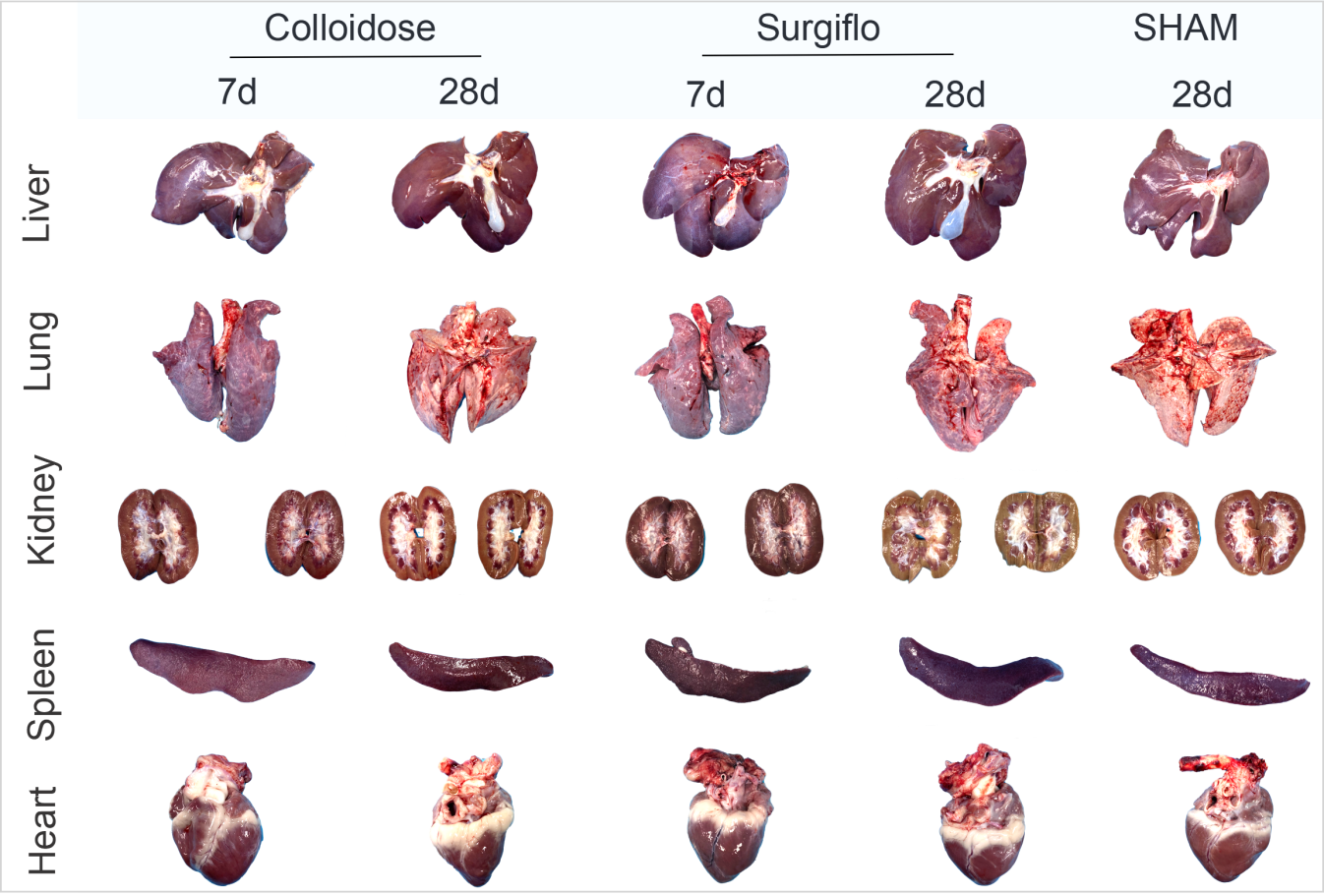

Figure S3. Representative images of major organs collected 28 days after surgery from Bama minipigs subjected to different treatment conditions. A standardized internal organ injury was created and treated with either Colloidose^®^ or Surgiflo™ flowable matrices. In the SHAM group, wounds were created without biomaterial implantation.

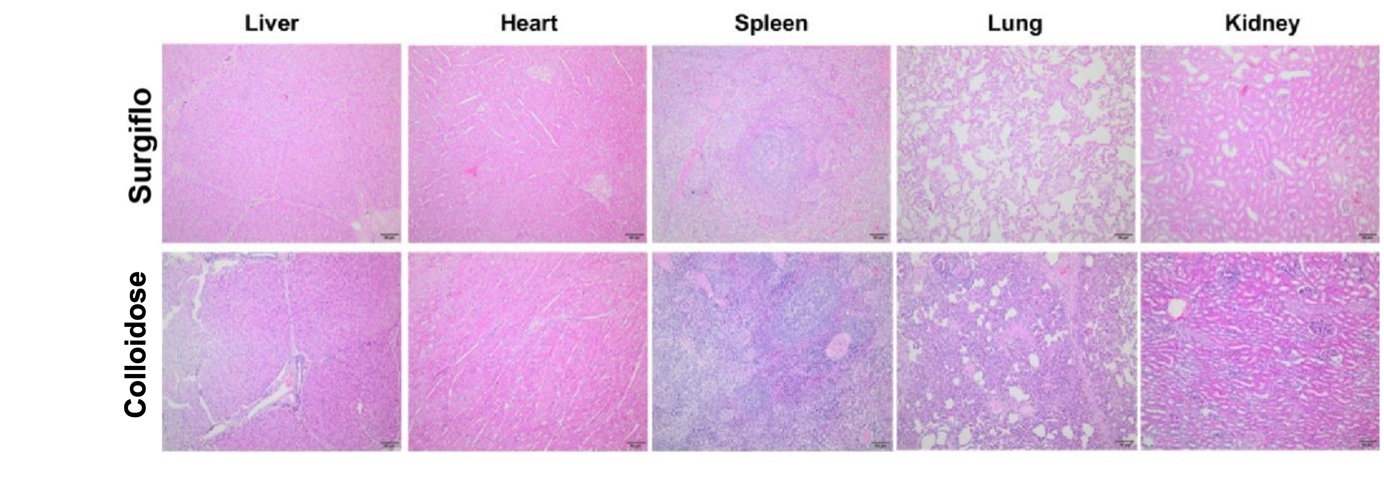

Figure S4. Histopathological analysis of non-surgical tissue from Bama minipigs in each group at one week post-operation. Tissue sections were collected from regions distant from the surgical site to evaluate potential systemic effects or off-target inflammatory responses.

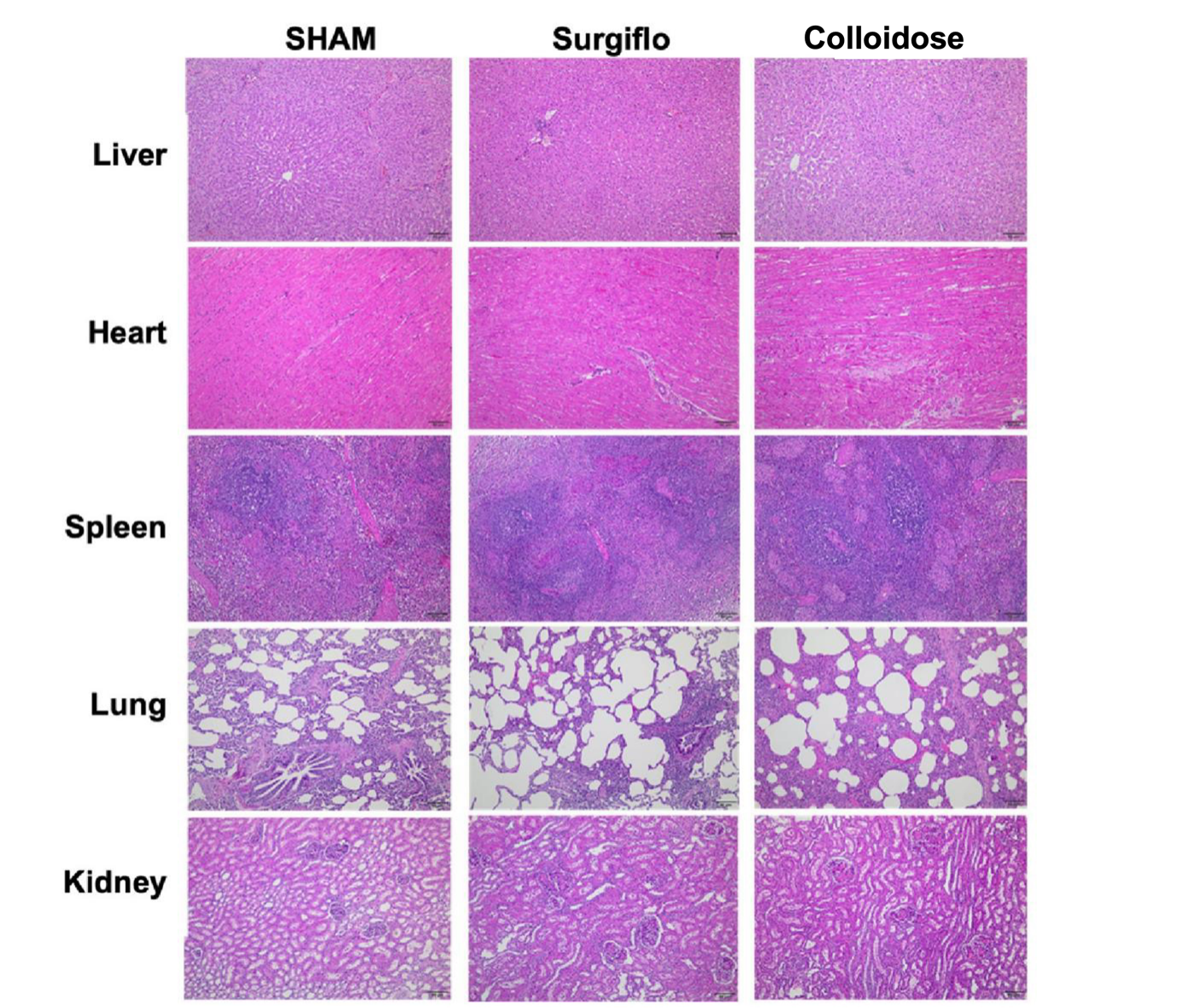

Figure S5. Histopathological analysis of non-surgical tissue from Bama minipigs in each group at 4 weeks post-operation. Tissue sections were harvested from areas distant from the implantation site to evaluate potential off-target effects. No significant histological differences were observed among treatment groups compared with the SHAM group.

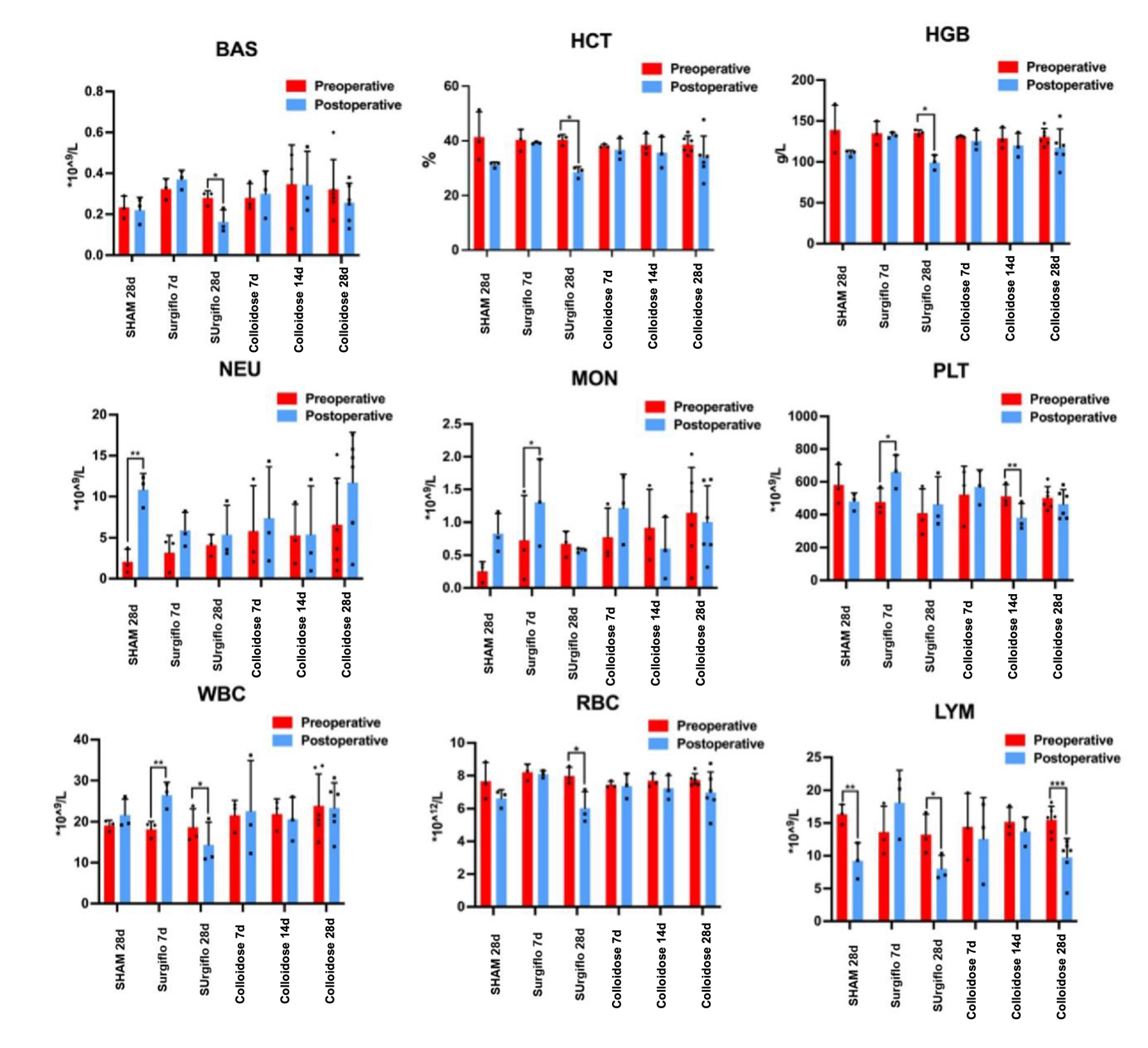

Figure S6. Routine blood test results in Bama minipigs before surgery (Preoperative, blue) and at necropsy (Postoperative, red) across treatment groups. Parameters include BAS (basophils), NEU (neutrophils), MON (monocytes), LYM (lymphocytes), WBC, RBC, HGB, HCT, and PLT (platelets). No significant changes were observed between pre- and post-operative values in any group, indicating good hematological biocompatibility of both Colloidose^®^ and Surgiflo™.

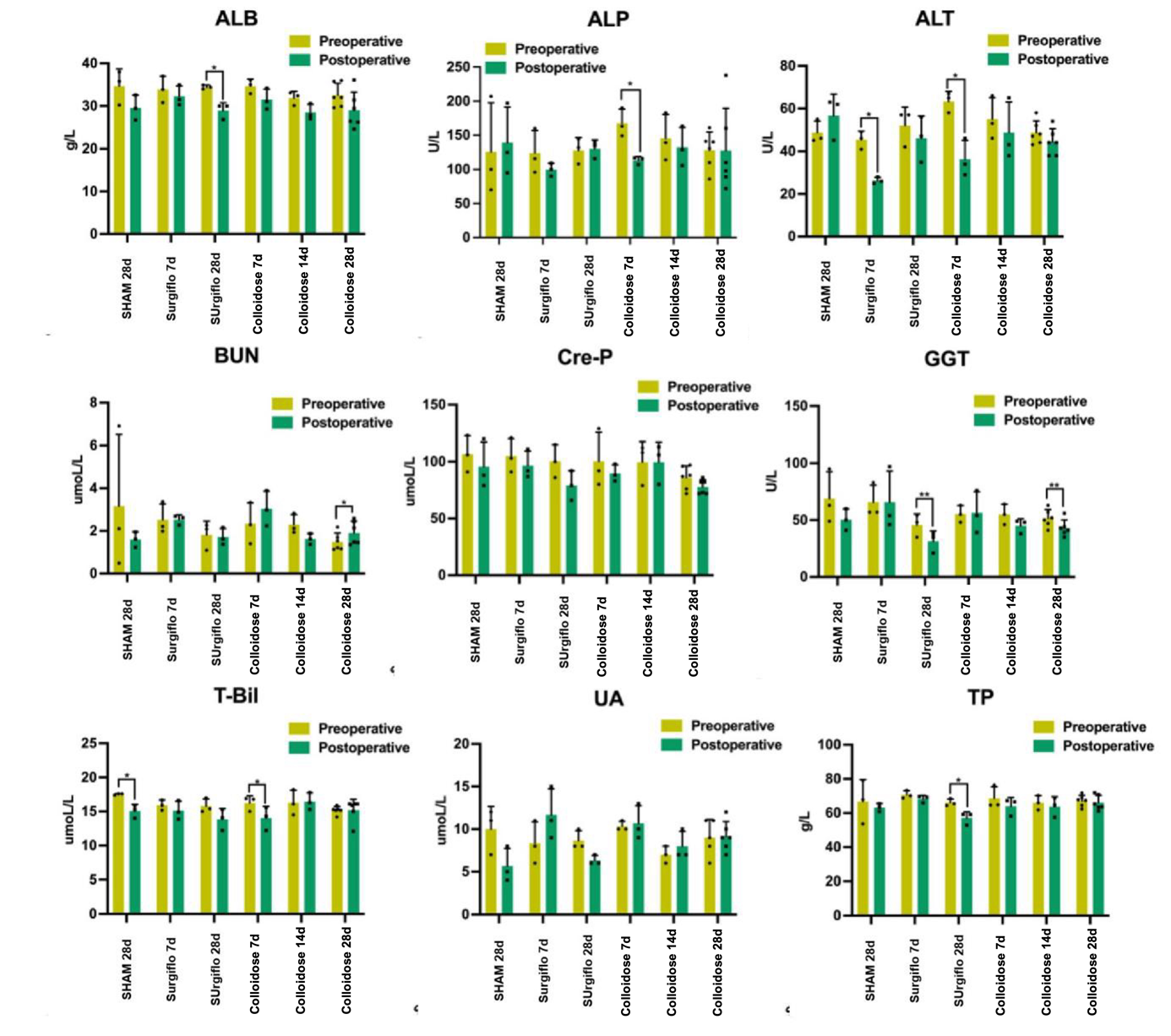

Figure S7.Blood biochemical test results in Bama minipigs before surgery (Preoperative, yellow) and at necropsy (Postoperative, green).Parameters include ALB, ALP, ALT, BUN, CRE-P, GGT, T-Bil, UA, and TP, representing liver and kidney function. No significant changes were observed between pre and post-operative values in any group, indicating that both Colloidose^®^ and Surgiflo™ did not induce systemic organ toxicity.
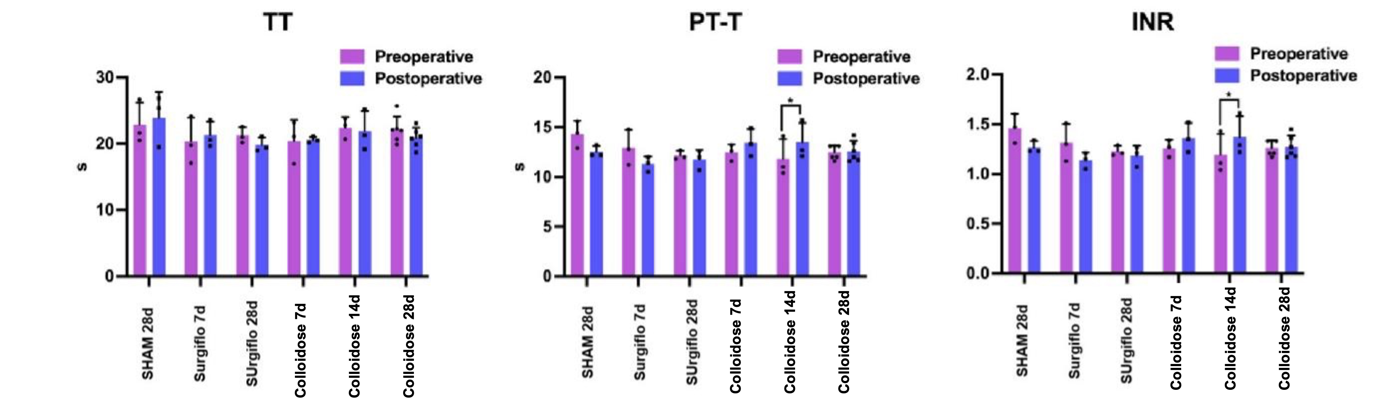

Figure S8.Comparison of results of coagulation analysis before and end the operation.

Table S1.Descriptive Statistics of Demographic Data of Subjects

| Project | | FAS | | | | |
| --- | --- | --- | --- | --- | --- | --- |
|  |  | Collodiose | Surgiflo | Testing method | Statistical measure | P Value |
| **age** |  |  |  |  |  |  |
| N(missing) |  | 174(0) | 174(0) | T-test | t=0.33 | 0.7447 |
| Mean±SD |  | 47.76±13.37 | 48.23±13.28 |  |  |  |
| median |  | 49 | 49 |  |  |  |
| Q1,Q3 |  | 37,58 | 40,58 |  |  |  |
| Min,Max |  | 18,74 | 18,4 |  |  |  |
| **Height** |  |  |  |  |  |  |
| N(missing) |  | 174(0) | 174(0) | T-test | t=0.74 | 0.4613 |
| Mean±SD |  | 163.51±8.33 | 162.89±7.34 |  |  |  |
| median |  | 162.5 | 162 |  |  |  |
| Q1,Q3 |  | 158,170 | 159,168 |  |  |  |
| Min,Max |  | 140,183 | 143,182 |  |  |  |
| **Weight** |  |  |  |  |  |  |
| N(missing) |  | 174(0) | 174(0) | T-test | t=0.27 | 0.7871 |
| Mean±SD |  | 66.48±13.12 | 66.84±11.41 |  |  |  |
| median |  | 64 | 66 |  |  |  |
| Q1,Q3 |  | 58,75 | 59,75 |  |  |  |
| Min,Max |  | 40,130 | 38,100 |  |  |  |
| **Body temperature** | |  |  |  |  |  |
| N(missing) |  | 174(0) | 174(0) | T-test | t=0.82 | 0.4108 |
| Mean±SD |  | 36.40±0.22 | 36.43±0.26 |  |  |  |
| median |  | 36.4 | 36.5 |  |  |  |
| Q1,Q3 |  | 36.2,36.5 | 36.3,36.5 |  |  |  |
| Min,Max |  | 36,37 | 35.5,37.2 |  |  |  |
| **Respiratory Rate (breaths per minute)** | | |  |  |  |  |
| N(missing) |  | 174(0) | 174(0) | T-test | t=2.15 | 0.0324 |
| Mean±SD |  | 18,89±1.01 | 19.12±1.03 |  |  |  |
| median |  | 19 | 19 |  |  |  |
| Q1,Q3 |  | 18,20 | 18,20 |  |  |  |
| Min,Max |  | 16,21 | 15,21 |  |  |  |
| **Diastolic Blood Pressure** | | |  |  |  |  |
| N(missing) |  | 172(2) | 172(2) | T-test | t=0.84 | 0.4278 |
| Mean±SD |  | 85.53±11.35 | 81.48±10.92 |  |  |  |
| median |  | 79 | 80 |  |  |  |
| Q1,Q3 |  | 74,88 | 73,89 |  |  |  |
| Min,Max |  | 55,115 | 57,116 |  |  |  |
| **Systolic Blood Pressure** | | |  |  |  |  |
| N(missing) |  | 172(2) | 172(2) | T-test | t=1.02 | 0.4033 |
| Mean±SD |  | 127.61±15.53 | 129.07±16.79 |  |  |  |
| median |  | 127 | 128 |  |  |  |
| Q1,Q3 |  | 117.5,135.3 | 118,139.50 |  |  |  |
| Min,Max |  | 87,178 | 90,183 |  |  |  |
| **Pulse Rate (beats per minute)** | | |  |  |  |  |
| Mean±SD |  | 79.38±10.55 | 78.29±9.28 | T-test | t=1.02 | 0.3085 |
| median |  | 78 | 78 |  |  |  |
| Q1,Q3 |  | 72,84 | 72,83 |  |  |  |
| Min,Max |  | 53,120 | 51,101 |  |  |  |
| **Gender** |  |  |  |  |  |  |
| Male |  | 59(33.91) | 49(28.16) | Chi-Square Test | 1.34 | 0.2466 |
| Female |  | 115(66.09) | 125(71.84) |  |  |  |
| Total |  | 174(100) | 174(100) |  |  |  |
| **Ethnicity** |  |  |  |  |  |  |
| Han Nationality | | 168(96.55) | 170(97.7) | Chi-Square Test | 0.41 | 0.521 |
| Other |  | 6(3.45) | 4(2.30) |  |  |  |
| Total |  | 174(100) | 174(100) |  |  |  |
| **Marital Status** | |  |  |  |  |  |
| Married |  | 157(90.23) | 159(91.38) | CMH test | 1.01 | 0.6036 |
| Unmarried |  | 14(8.05) | 14(8.05) |  |  |  |
| Other |  | 3(1.72) | 1(0.57) |  |  |  |
| Total |  | 174(100) | 174(100) |  |  |  |

Table S2. Hemostatic efficacy within 5 minutes in different analysis populations. FAS (Full Analysis Set): all animals included according to the intention-to-treat principle. PPS (Per Protocol Set): animals that completed the study without major protocol deviations.

| Project | | FAS | | PPS | |
| --- | --- | --- | --- | --- | --- |
|  |  | Colloidose | Surgiflo | Colloidose | Surgiflo |
| **Hemostasis efficacy rate within 5 minutes** | | | |  |  |
| Effective |  | 174(100.00) | 174(100.00) | 174(100.00) | 172(100.00) |
| Ineffective |  | 0(0.00) | 0(0) | 0(0) | 0(0) |
| Total |  | 174(100.00) | 174(100.00) | 174(100.00) | 172(100.00) |
| **Comparison of therapeutic effects between two groups** | | | | |  |
| Statistical measure | | Z=0.00 | | Z=0.00 | |
| P value |  | 1 | | 1 | |
| **95% confidence interval for the difference in effectiveness between two groups** | | | | | |
| Newcombe-Wilson method | | (-0.0216,0.0216) | | (-0.0216,0.0218) | |
| Haldane method | | (0,0) | | (0,0.0001) | |
| Jeffreys-Perks method | | (-0.0112,0.0112) | | (-0.0112,0.0113) | |
| Brown-Li method | | (-0.0112,0.0112) | | (-0.0112,0.0113) | |

Table S3. Hemostatic efficacy within 3 minutes in different analysis populations. FAS (Full Analysis Set): all animals included according to the intention-to-treat principle. PPS (Per Protocol Set): animals that completed the study without major protocol deviations.

| Project | | FAS | | PPS | |
| --- | --- | --- | --- | --- | --- |
|  |  | Colloidose | Surgiflo | Colloidose | Surgiflo |
| **Hemostasis efficacy rate within 3 minutes** | | | |  |  |
| Effective |  | 172(98.85) | 165(94.83) | 172(98.85) | 163(94.77) |
| Ineffective |  | 2(1.15) | 9(5.17) | 2(1.15) | 9(5.23) |
| Total |  | 174(100.00) | 174(100.00) | 174(100.00) | 172(100.00) |
| **Comparison of therapeutic effects between two groups** | | | | |  |
| Statistical measure | | Z=2.14 | | Z=2.16 | |
| P value |  | 0.0324 | | 0.0308 | |
| **95% confidence interval for the difference in effectiveness between two groups** | | | | | |
| Newcombe-Wilson method | | (0.0021,0.0847) | | (0.0025,0.0857) | |
| Haldane method | | (0.0035,0.0761) | | (0.0038,0.0771) | |
| Jeffreys-Perks method | | (0.002,0.0776) | | (0.0023,0.0786) | |
| Brown-Li method | | (0.002,0.078) | | (0.0023,0.079) | |

Table S4. Routine hematological parameters before and after treatment with Colloidose® or Surgiflo™.

| Project | Screening period normal | | Abnormal during the screening period | | Missing data situation | | | Total |
| --- | --- | --- | --- | --- | --- | --- | --- | --- |
|  | Normal after treatment | Abnormal after treatment | Normal after treatment | Abnormal after treatment | Pre-treatment missing | Post-treatment missing | Missing before and after treatment |  |
| WBC |  |  |  |  |  |  |  |  |
| Colloidose | 127 | 26 | 14 | 5 | 0 | 2 | 0 | 174 |
| Surgiflo | 129 | 38 | 5 | 1 | 1 | 0 | 0 | 174 |
| RBC |  |  |  |  |  |  |  |  |
| Colloidose | 93 | 57 | 3 | 19 | 0 | 2 | 0 | 174 |
| Surgiflo | 94 | 55 | 3 | 21 | 1 | 0 | 0 | 174 |
| PLT |  |  |  |  |  |  |  |  |
| Colloidose | 143 | 9 | 9 | 11 | 0 | 2 | 0 | 174 |
| Surgiflo | 150 | 6 | 8 | 9 | 1 | 0 | 0 | 174 |
| LYM |  |  |  |  |  |  |  |  |
| Colloidose | 87 | 57 | 6 | 22 | 0 | 2 | 0 | 174 |
| Surgiflo | 86 | 65 | 10 | 12 | 1 | 0 | 0 | 174 |
| NEU |  |  |  |  |  |  |  |  |
| Colloidose | 109 | 41 | 13 | 9 | 0 | 2 | 0 | 174 |
| Surgiflo | 108 | 50 | 11 | 4 | 1 | 0 | 0 | 174 |
| CRP |  |  |  |  |  |  |  |  |
| Colloidose | 42 | 93 | 3 | 31 | 1 | 4 | 0 | 174 |
| Surgiflo | 38 | 102 | 1 | 28 | 4 | 1 | 0 | 174 |
